# PDA Closure Rates and Safety after Repeated Courses of Cyclooxygenase Inhibitors in Preterm Infants: A Systematic Review and Meta-Analysis

**DOI:** 10.64898/2026.09.09.26360534

**Authors:** Junichi Saito, Yuka Sano Wada, Kota Yoneda, Kunio Ogawa, Nobuhiko Kan, Tetsuya Isayama, Katsuaki Toyoshima

## Abstract

**Background:** Cyclooxygenase (COX) inhibitors have been used to treat hemodynamically significant patent ductus arteriosus (PDA) in preterm infants. If the PDA remains hemodynamically significant after an initial course, additional courses of COX inhibitors or surgical intervention are considered. This systematic review evaluates PDA closure rates and safety after repeated courses of COX inhibitors in preterm infants.

**Methods:** Embase, Medline, and two other databases were searched through April 2026 (pre-registered in PROSPERO [CRD42023454003]). Studies reporting PDA closure rates after two or more courses of COX inhibitors were included. Risk of bias was assessed using Joanna Briggs Institute tools. Meta-analyses with random-effects models were performed to calculate the proportions of PDA closure. Multilevel mixed-effects meta-regression was used to compare closure rates across treatment courses while accounting for within-study correlations.

**Results:** A total of 33 studies were included. The PDA closure rates after the first, second, and third courses of COX inhibitors were 62% (95% confidence interval [CI] 57–67%), 46% (95% CI 40–52%; *p* < 0.001 vs the first course), and 38% (95% CI 27–51%; *p* = 0.001 vs the first course), respectively. Lower closure rates with repeated courses were also observed across most subgroups (extremely preterm/extremely low birth weight, ibuprofen, indomethacin, and intravenous administration), except for the second course of oral administration. Data on adverse events were limited, preventing meta-analysis.

**Conclusions:** PDA closure rates after the second and third courses of COX inhibitors were lower than those after the first course in preterm infants. Evidence regarding the safety of repeated courses remains limited.

**Impact:**

- This systematic review is the first study to integrate the PDA closure rates in preterm infants following the second and third courses of COX inhibitor treatment.
- The PDA closure rate after the first course of COX inhibitor treatment was 62%, whereas those after the second and third courses were 46% and 38%, respectively.
- Multilevel meta-regression and pairwise comparisons showed that later COX inhibitor courses yielded significantly lower PDA closure rates than the first.
- Evidence with adverse events after repeated COX inhibitor treatment was limited and required further investigation.

## INTRODUCTION

The ductus arteriosus (DA), a fetal vascular shunt connecting the pulmonary artery and aorta, normally closes after birth. Failure of DA closure (i.e., patent DA [PDA]) is observed in preterm infants, with a prevalence exceeding 50% among infants born at less than 28 weeks’ gestation [1]. PDA causes systemic ischemia and pulmonary vascular congestion, and thus is associated with renal dysfunction, intestinal perforation, prolonged mechanical ventilation, and the development of chronic lung disease [2].

Management of PDA involves a choice among expectant management [3, 4], pharmacological treatment, and surgical intervention. The standard pharmacological treatment for PDA is cyclooxygenase (COX) inhibitors [5]. The COX inhibitors approved by the Food and Drug Administration for PDA are ibuprofen and indomethacin. When a persistent PDA continues to compromise an infant’s clinical condition after the initial course of COX inhibitor treatment, clinicians must weigh repeated pharmacological treatment against surgical intervention (e.g., ligation). Repeated courses of COX inhibitors may avoid the risks associated with surgery but may also increase the risk of adverse events, including renal dysfunction, gastrointestinal bleeding, necrotizing enterocolitis (NEC), and spontaneous intestinal perforation [5]. Therefore, evidence regarding the actual PDA closure rate and safety after repeated courses of COX inhibitors is essential for guiding treatment decisions. This review systematically summarizes the best available evidence on these outcomes for preterm infants.

## METHODS

This systematic review and meta-analysis were conducted and reported in accordance with the Preferred Reporting Items for Systematic Reviews and Meta-Analyses (PRISMA) guidelines (PRISMA checklists in Tables S1 and S2) [6]. The protocol was pre-registered with the International Prospective Register of Systematic Reviews (PROSPERO: CRD42023454003).

### Eligibility Criteria

This systematic review included studies reporting PDA closure rates after two or more courses of COX inhibitor treatment in preterm infants with hemodynamically significant PDA. COX inhibitors included both ibuprofen and indomethacin. We included randomized controlled trials (RCTs), non-RCTs, interrupted time series studies, cohort studies, case-control studies, before-and-after studies, and case series with 10 or more cases. We excluded studies involving only term infants; studies evaluating acetaminophen alone; studies using COX inhibitors to prevent intraventricular hemorrhage; case reports; studies based solely on proceedings; and animal studies. Outcome data for the first, second, and third courses were extracted from the included studies. The primary outcome was PDA closure rates after each course of COX inhibitor treatment. Secondary outcomes were COX inhibitor-related adverse events, including renal impairment and gastrointestinal complications, after each treatment course.

### Search Strategy

A dedicated literature search specialist (K.O.) conducted electronic searches on August 10, 2023, and updated the searches on April 9, 2026, in MEDLINE, EMBASE, the Cochrane Central Register of Controlled Trials, and Cumulative Index to Nursing and Allied Health Literature (CINAHL). The search strategy used the terms “preterm infant” AND “patent ductus arteriosus” AND “cyclooxygenase inhibitor.” Database-specific search strategies are detailed in Table S3. No restrictions on language or publication date were applied.

### Data Extraction and Risk of Bias Assessment

Rayyan software was used for study screening. Titles and abstracts were independently screened by two authors (J.S. and YS.W.), with disagreements resolved by consensus through discussion. The same authors independently reviewed the full texts of potentially eligible studies. After full-text review, descriptive data (authors, publication year, country, sample size, gestational age, birth weight, sex, type of COX inhibitor, and route of administration) and outcome data (PDA closure, renal dysfunction, gastrointestinal complications, and other drug-related complications) were extracted. PDA closure was defined as clinical resolution (i.e., no need for additional PDA-directed treatment) to account for variation among the included studies in the definition of closure (anatomical or clinical) and the timing of echocardiographic assessment. Risk of bias was assessed using the Joanna Briggs Institute (JBI) Critical Appraisal Checklist for Case Series [7].

### Data Synthesis and Statistical Analysis

Pooled PDA closure rates and their 95% confidence intervals (CIs) after the first, second, and third courses were calculated using an inverse-variance weighted random-effects model using R software (version 4.6.0) with the metafor package (version 8.5-0) [8, 9]. Heterogeneity between studies was assessed visually using forest plots and quantitatively using Cochran’s Q test and the *I*² statistic. An *I*² value < 30% indicates low heterogeneity, while > 50% denotes high heterogeneity [10].

Outcome data were analyzed for all preterm infants and for predefined subgroups. The predefined subgroups included infants with extreme prematurity (EPT; gestational age < 28 weeks) or extremely low birth weight (ELBW; birth weight < 1000 g), referred to as EPT/ELBW infants; infants treated with ibuprofen; infants treated with indomethacin; infants who received intravenous administration; and infants who received oral administration. We evaluated trends in PDA closure rates across treatment courses using a multilevel mixed-effects meta-regression model. To account for potential within-study correlation among course-specific estimates, we incorporated study-level random intercepts and modeled treatment course as a continuous variable. Additionally, we conducted pairwise comparisons to test for significant differences in PDA closure rates between specific treatment courses (first vs second, first vs third, and second vs third) using paired data from the same studies. Subgroup comparisons were also performed within each treatment course to evaluate differences between (1) EPT/ELBW infants and preterm infants from cohorts that did not overlap with the EPT/ELBW subgroup, (2) ibuprofen and indomethacin, and (3) intravenous and oral administration.

Publication bias was assessed by funnel plots and Egger’s regression test. Statistical significance was set at *p* < 0.05. The overall certainty of evidence was assessed using the Grading of Recommendations Assessment, Development, and Evaluation (GRADE) approach [11].

## RESULTS

### Results of Literature Search and Screening

Database searches identified 4,117 records (Fig. 1). After duplicate records were removed through automated and manual processes, 2,904 records underwent title and abstract screening. Of these, 2,712 records were excluded, leaving 192 records for full-text assessment. After full-text review, 159 records were excluded (Fig. 1), and 33 studies [12–44] were included in the meta-analyses. All included studies were published in peer-reviewed journals. A review of the reference lists of the included studies identified no additional eligible studies.

**Figure 1.**
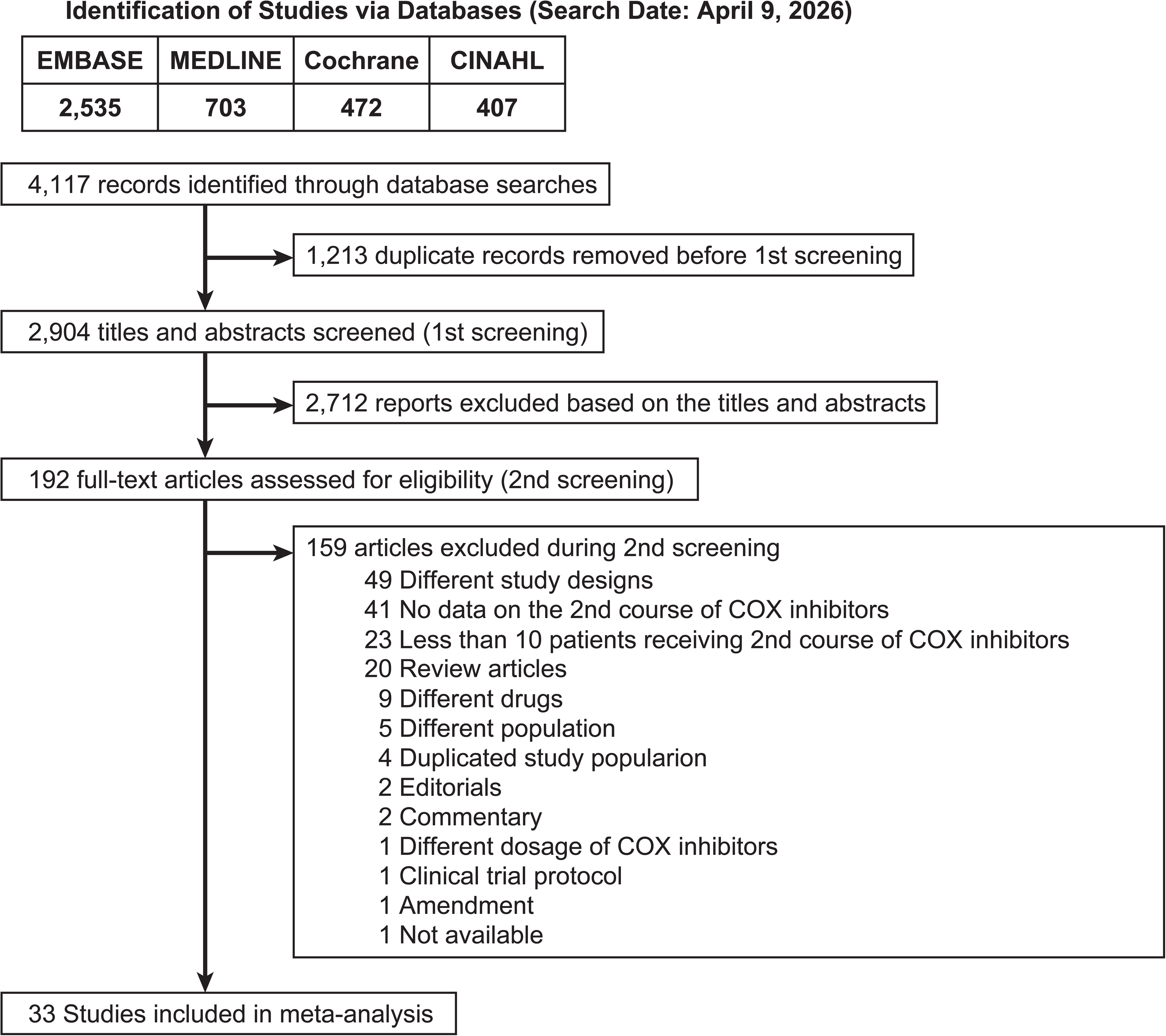
Flow diagram of the study selection process.

### Basic Characteristics of Included Literature

Characteristics of the included studies are presented in Table 1 (details in Table S4). The studies were published between 2002 and 2026 and were conducted in Italy [12, 17, 29, 34, 40, 42], Turkey [18, 22, 31, 33, 41], the United States [14, 19, 21, 44], Canada [24, 26, 36], Egypt [20, 32, 35], South Korea [27, 39], Singapore [13, 15], the United Kingdom [16], the Netherlands [25], Albania [30], India [37], China [43], Hong Kong [28], Taiwan [23], and Iran [38]. Among the 33 included studies, a total of 4,211 infants received COX inhibitors. In all studies, COX inhibitors were administered after PDA was confirmed by ultrasound examination and clinical symptoms attributable to PDA were identified. Four studies reported the use of indomethacin, 20 reported the use of ibuprofen, and 9 reported the use of either indomethacin or ibuprofen. Intravenous administration was reported in 14 studies, oral administration in 7 studies, and either intravenous or oral administration in 7 studies; the route of administration was not specified in 5 studies. Ten studies administered a third course of COX inhibitors to 10 or more patients [14, 17, 19, 21, 24, 25, 29, 33, 36, 41]. Only three studies reported adverse events after each course of repeated COX inhibitor administration [14, 19, 20].

**Table 1.**
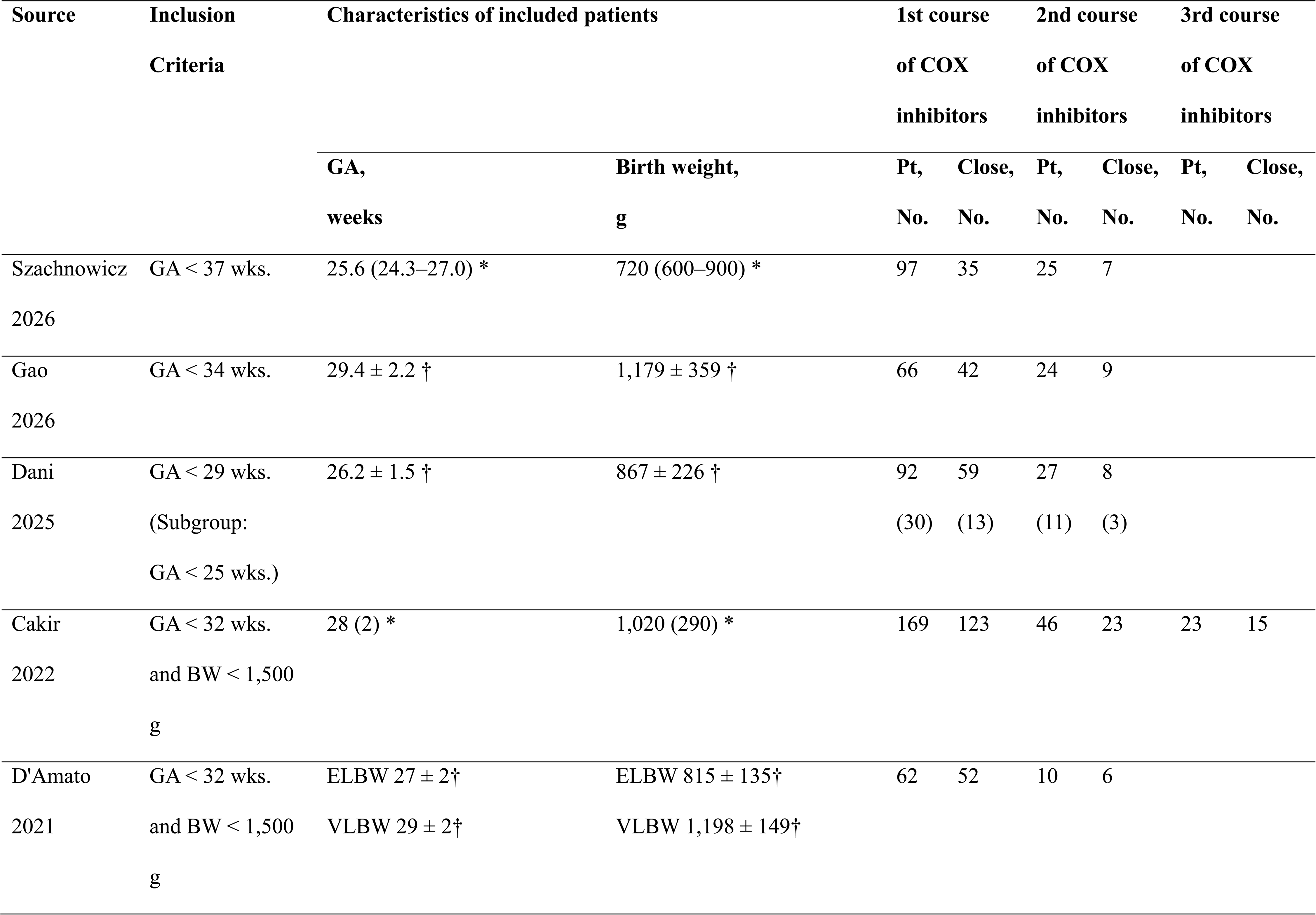
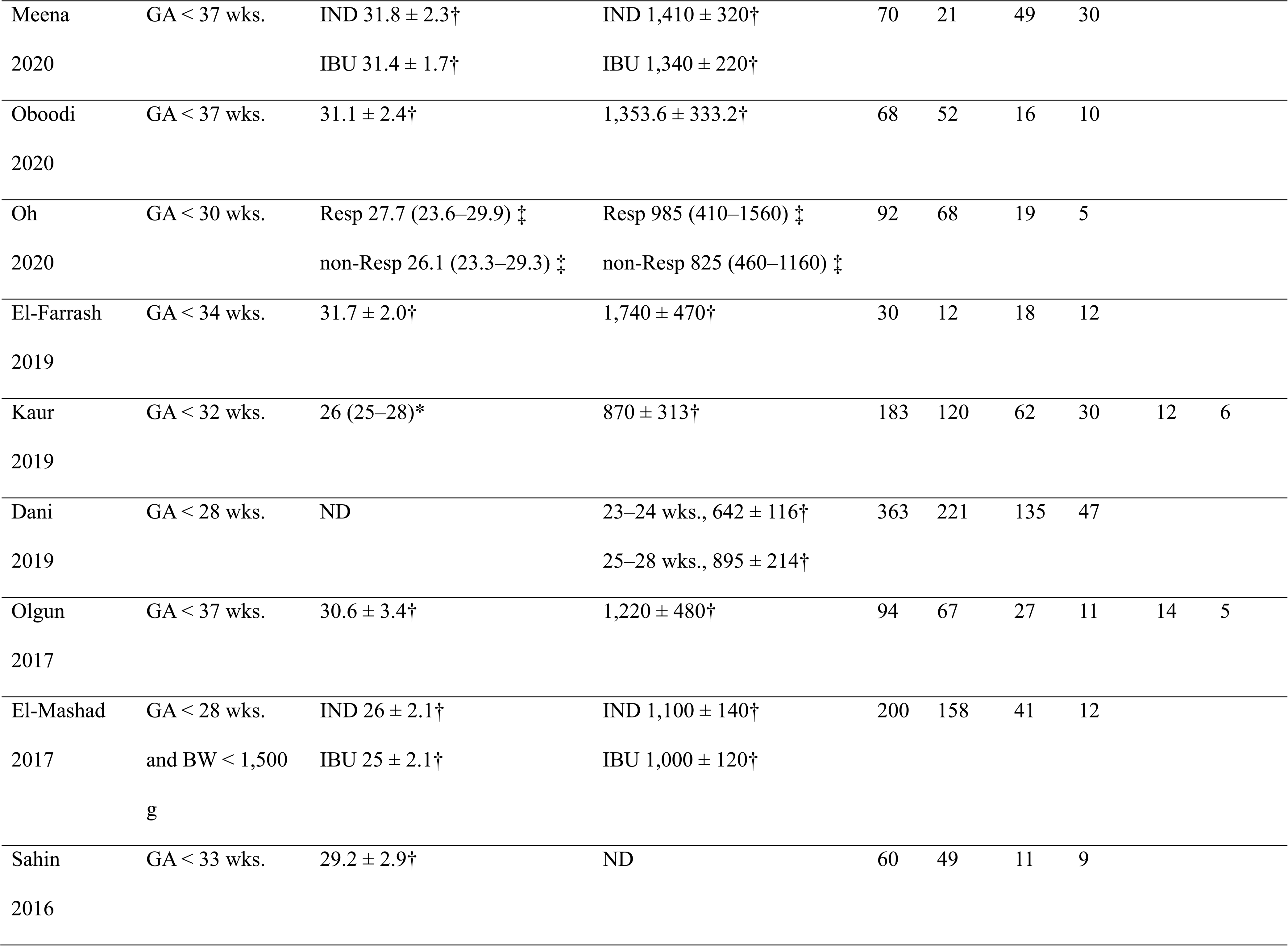
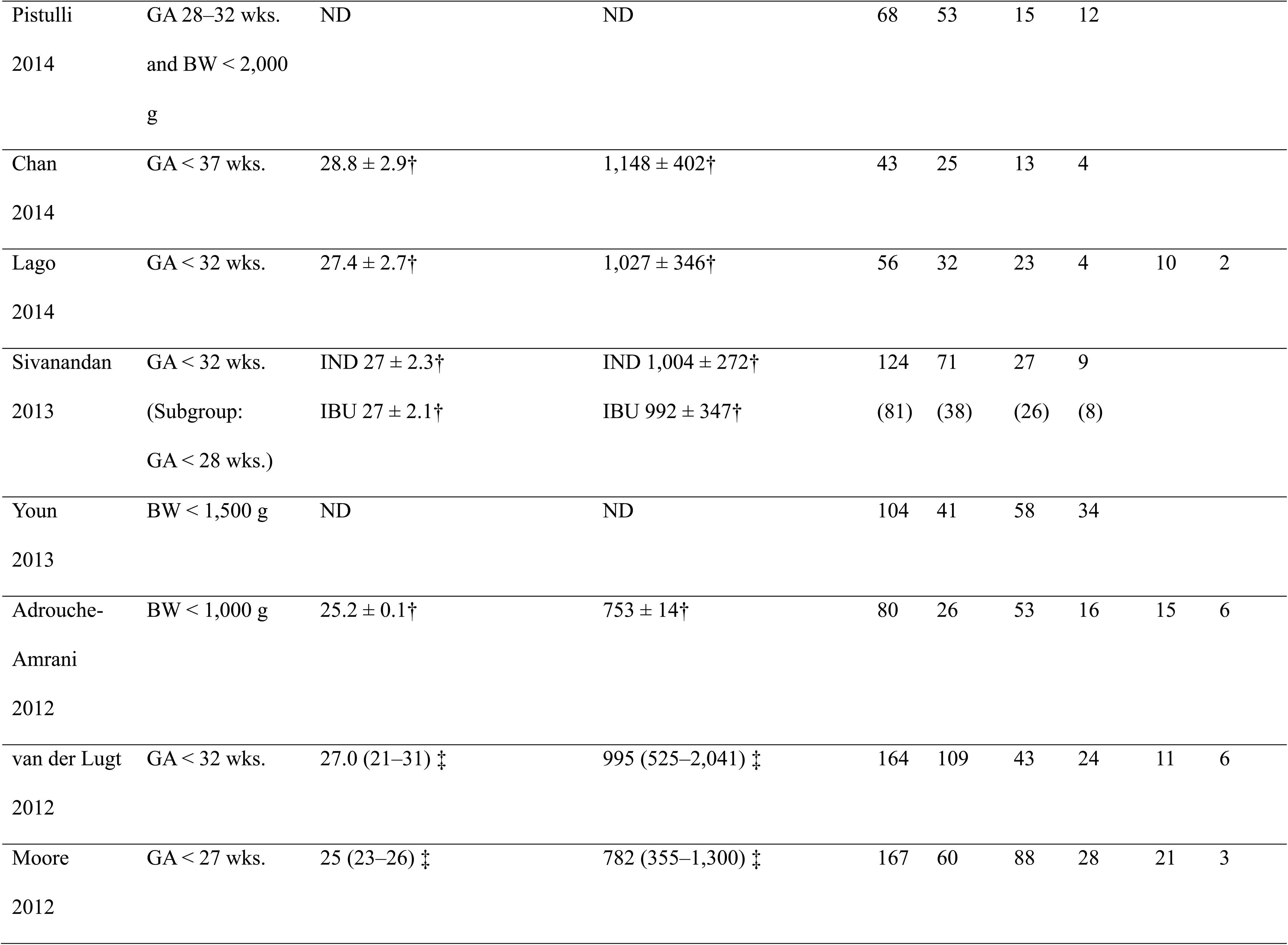
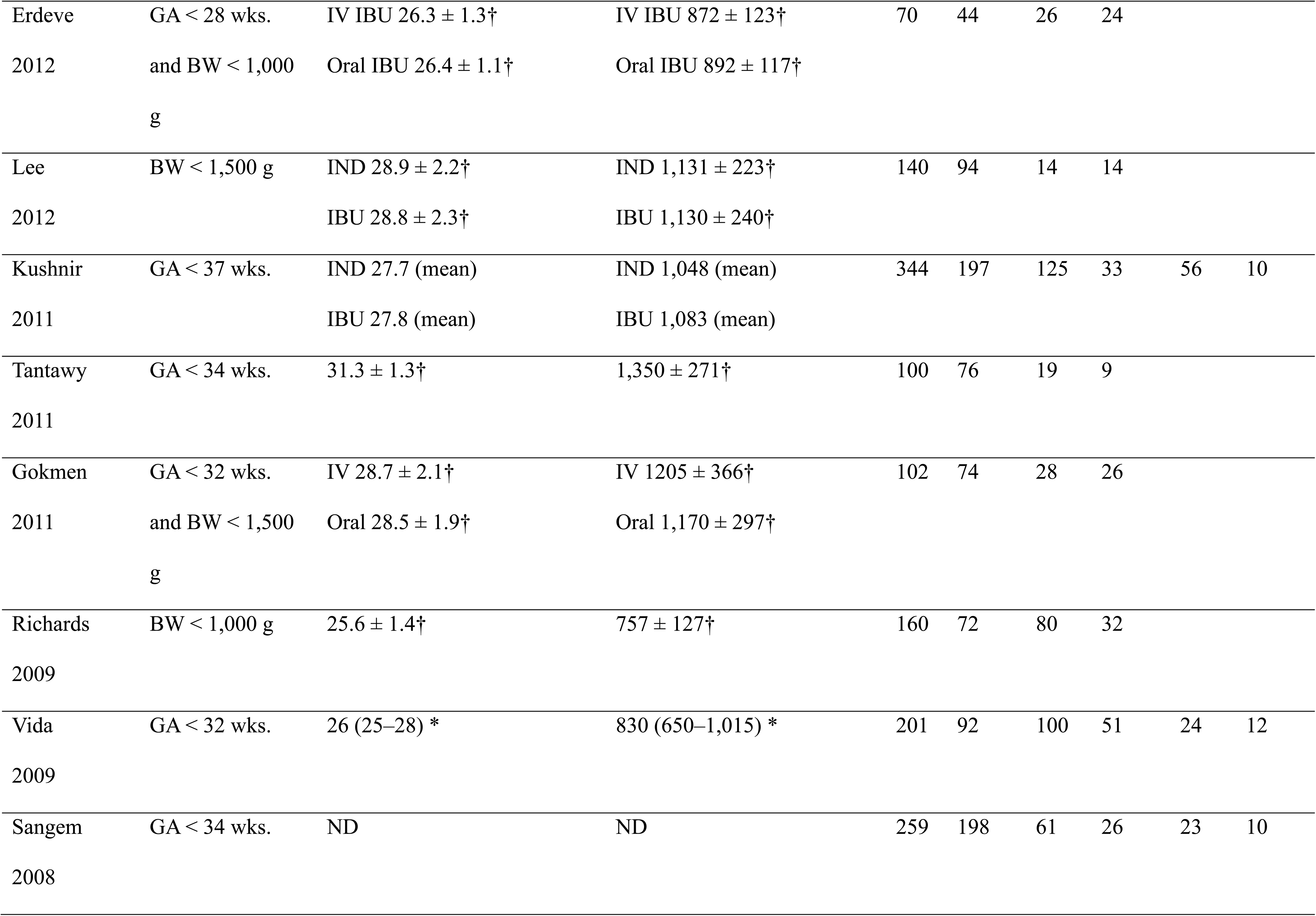
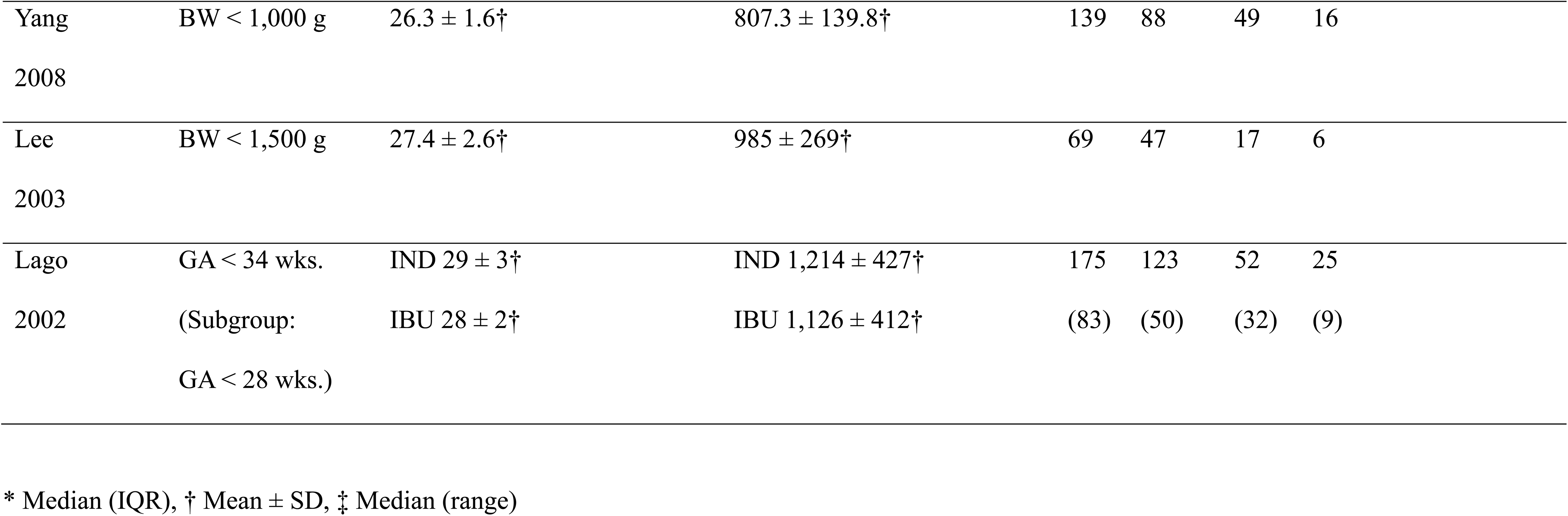
Basic characteristics of the 33 included studies.

### Results of Literature Quality Evaluation

The risk of bias assessment using the JBI checklist is provided in Table S5. According to the JBI checklist, 11 of the 33 studies had a low risk of bias [13, 14, 16, 17, 21, 24–26, 33, 38, 42]. Twenty-one studies had a serious risk of bias due to unclear reporting of clinical information, including participants’ characteristics and outcomes [12, 15, 18–20, 22, 23, 27–32, 34–36, 39–41, 43, 44]. One study had a very serious risk of bias because the study period was insufficiently described [37].

### Primary Outcome Measurement: PDA closure rate

Results for the first and second courses were available from the 33 studies [12–44], whereas third-course results were available from only 10 studies [14, 17, 19, 21, 24, 25, 29, 33, 36, 41]. Meta-analyses of 33 studies [12–44] calculated a PDA closure rate of 62% (2,601/4,211; 95% CI, 57–67%) after the first course of COX inhibitors, with substantial heterogeneity (*I²* = 89.7%, *p* < 0.0001; Fig. 2a). The calculated closure rates after the second and third courses were 46% (612/1,401; 95% CI, 40–52%; *I²* = 73.1%; *p* < 0.0001; Fig. 2b) and 38% (75/209; 95% CI, 27–51%; *I²* = 63.6%; *p* = 0.0033; Fig. 2c), respectively. Multilevel meta-regression analysis showed that a greater number of COX inhibitor courses was associated with lower PDA closure rates (odds ratio, 0.56; 95% CI, 0.50–0.62; *p* < 0.001; Table S6). Pairwise comparisons demonstrated significantly lower closure rates after the second and third courses than after the first course (1st vs 2nd, *p* < 0.001; 1st vs 3rd, *p* < 0.001; Table S7). Figure 3 illustrates the PDA closure rates after each COX inhibitor course in the overall preterm infant population.

**Figure 2.**
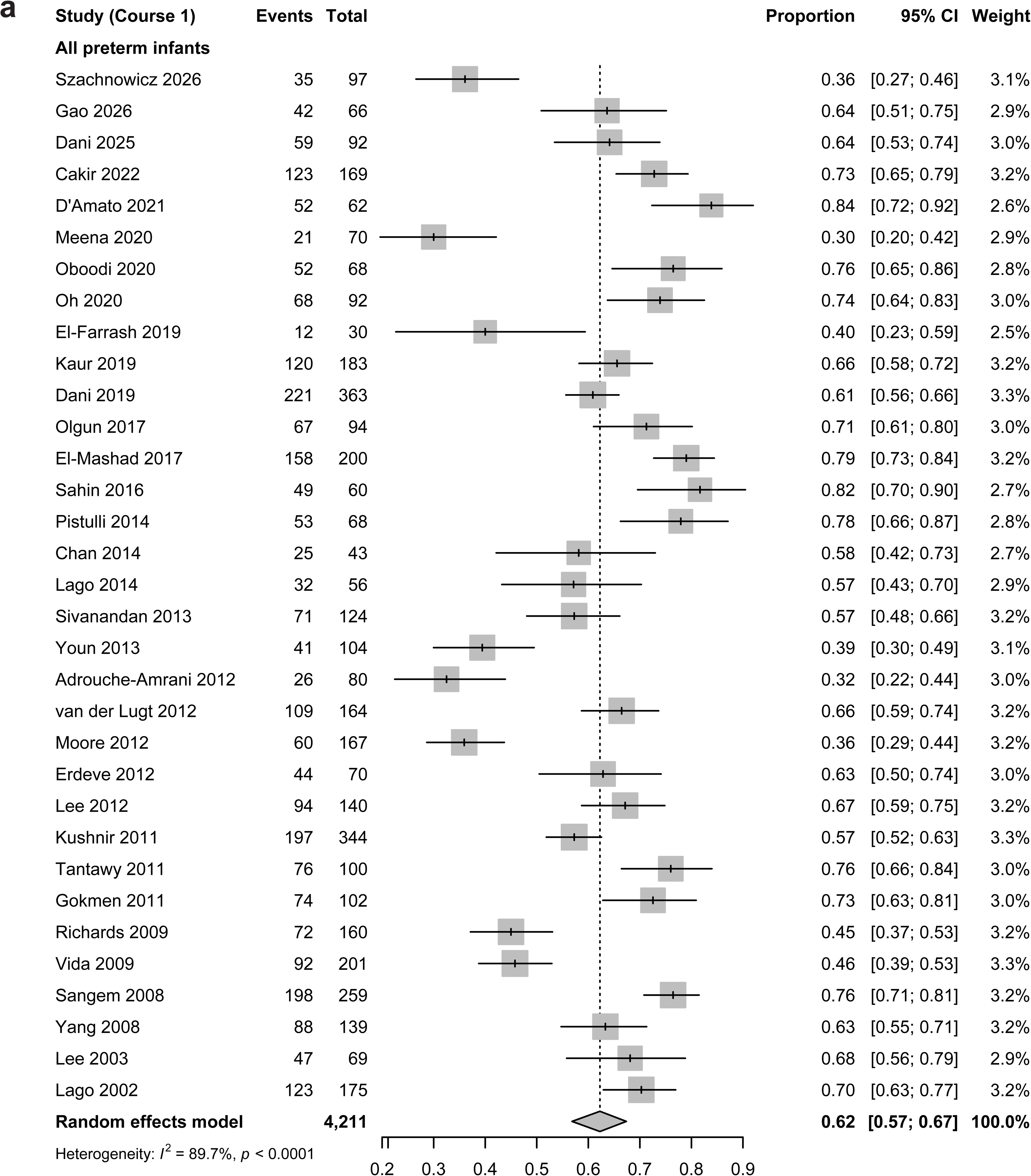
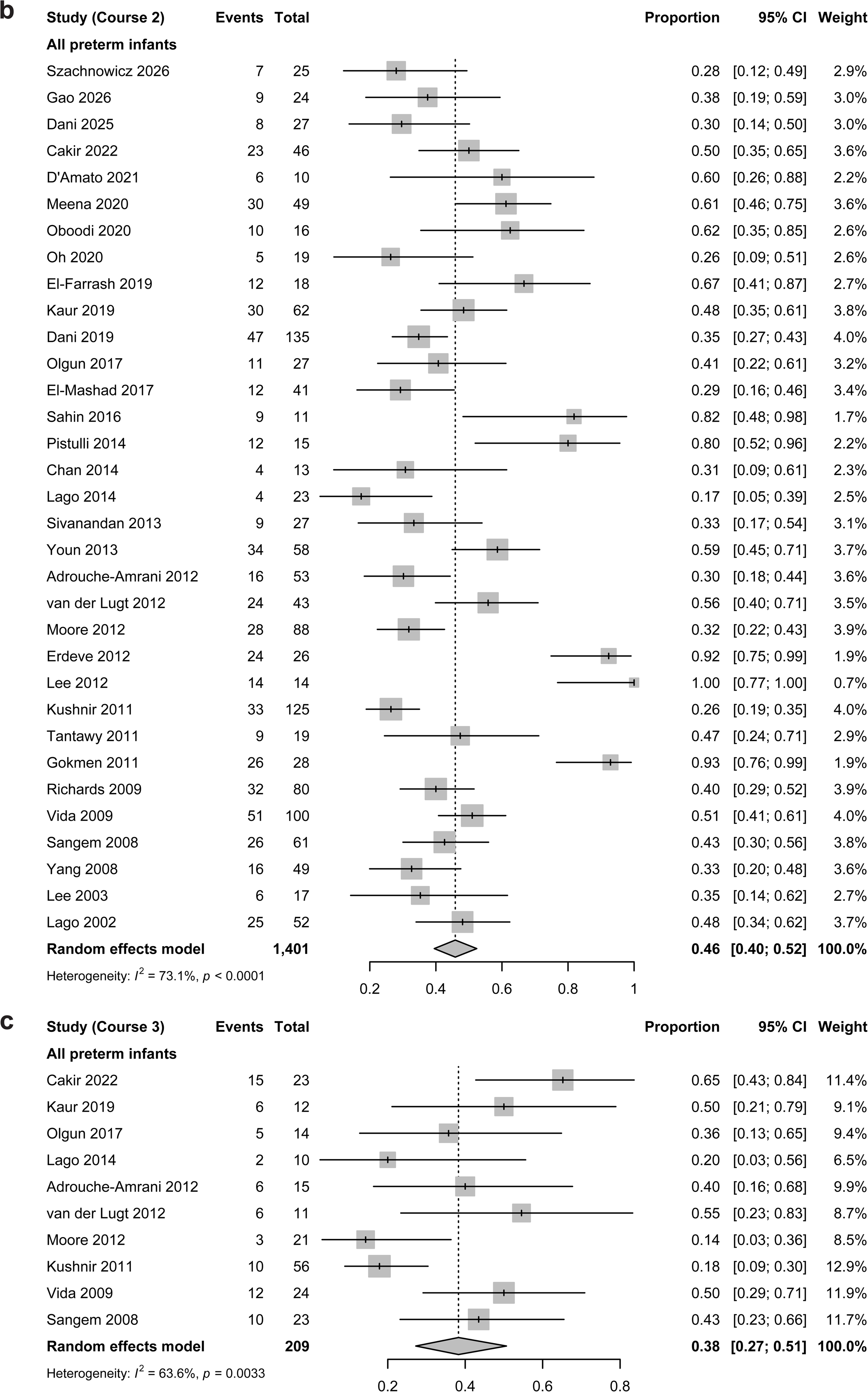
Forest plots of PDA closure rates after COX inhibitor treatment in the overall preterm infant population. **a.** Closure rate after the first course. **b.** Closure rate after the second course. **c.** Closure rate after the third course.

**Figure 3.**
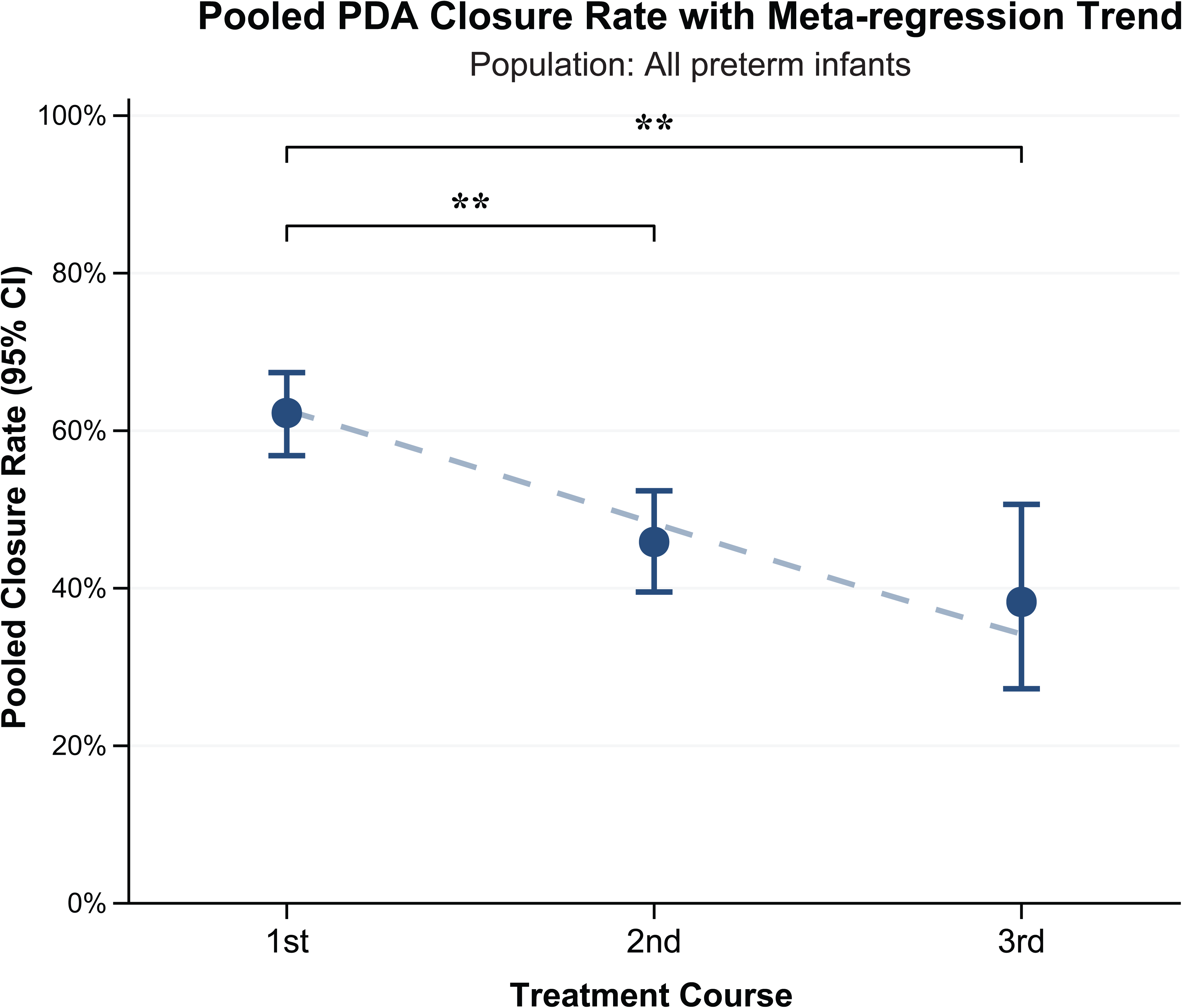
Pooled PDA closure rates and meta-regression trends in the overall preterm infant population. Pooled PDA closure rates are shown after the first, second, and third treatment courses, with corresponding 95% confidence intervals (CIs). Meta-regression analysis illustrates temporal trend in PDA closure rates across treatment courses.

### Subgroup Analysis

To assess potential sources of the substantial heterogeneity observed in the meta-analysis, we performed pre-registered subgroup analyses.

#### 1. Gestational Age and Birth Weight

Among EPT/ELBW infants, the pooled PDA closure rates after the first, second, and third courses of COX inhibitors were 52% (95% CI, 41–63%), 33% (95% CI, 29–37%), and 26% (95% CI, 8–57%), respectively (Fig. 4). Substantial heterogeneity persisted across courses in EPT/ELBW infants (*I²* = 55.4–92.4%; Fig. 4). Multilevel meta-regression analysis showed that a greater number of COX inhibitor courses was associated with lower PDA closure rates (odds ratio, 0.50; 95% CI, 0.41–0.61; *p* < 0.001; Table S6), and pairwise comparison demonstrated a significantly lower closure rate after the second course than after the first course (1st vs 2nd, *p* = 0.043; Table S7). Figure 5 illustrates PDA closure rates after each course of COX inhibitors in EPT/ELBW infants. To compare EPT/ELBW infants with other preterm infants, we excluded cohorts with overlapping EPT/ELBW populations to ensure independent comparison groups. PDA closure rates were consistently lower among EPT/ELBW infants than among preterm infants from non-overlapping cohorts (Table S8).

**Figure 4.**
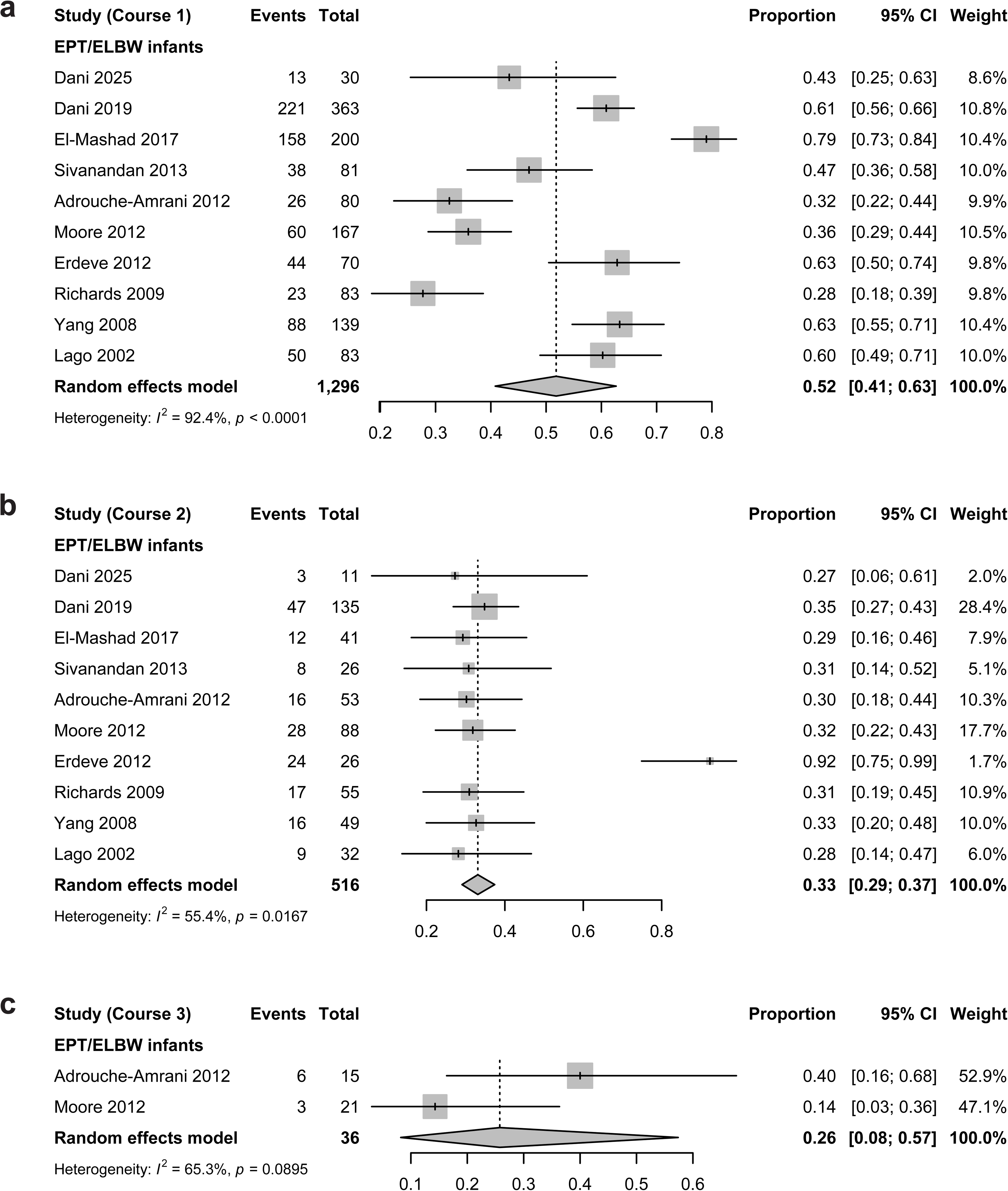
Forest plots of PDA closure rates after COX inhibitor treatment in EPT/ELBW infants. **a.** Closure rate after the first course. **b.** Closure rate after the second course. **c.** Closure rate after the third course.

**Figure 5.**
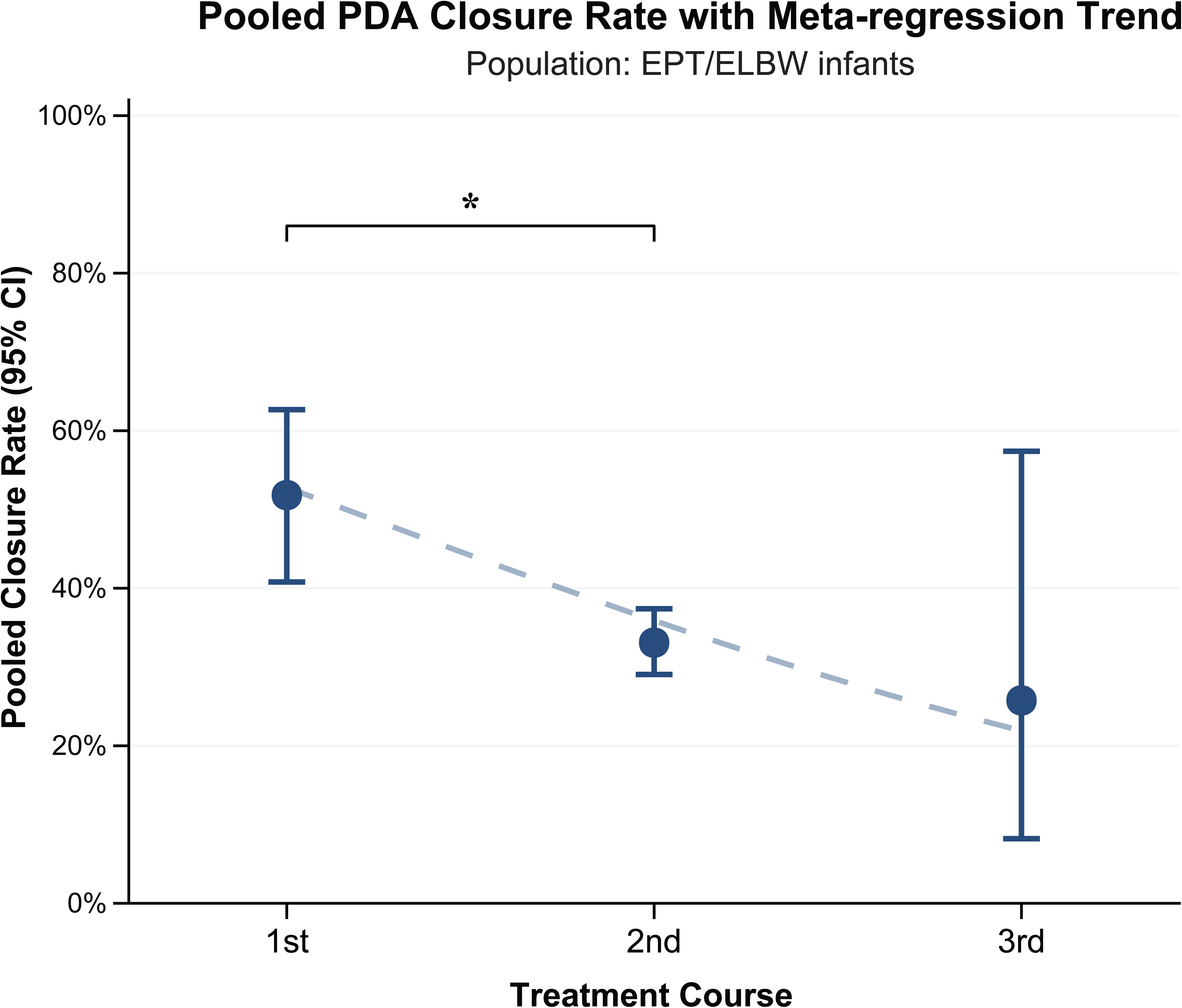
Pooled PDA closure rates and meta-regression trends in EPT/ELBW infants. Pooled PDA closure rates are shown after the first, second, and third treatment courses, with corresponding 95% confidence intervals (CIs). Meta-regression analysis illustrates temporal trends in PDA closure rates across treatment courses.

#### 2. Type of Drug (Ibuprofen or Indomethacin)

For both ibuprofen and indomethacin, PDA closure rates decreased from the first to the third course (ibuprofen: 63%, 49%, and 40%; indomethacin: 60%, 38%, and 24%; Figs. S1–S4), with substantial heterogeneity remaining across these analyses (*I²* = 45.4–92.0%; Figs. S1 and S3). Multilevel meta-regression confirmed that a greater number of treatment courses was associated with lower PDA closure rates for both medications (ibuprofen: odds ratio 0.50; 95% CI, 0.41– 0.61; *p* < 0.001; indomethacin: odds ratio 0.44; 95% CI, 0.36–0.54; *p* < 0.001; Table S6). There were no statistically significant differences in PDA closure rates between ibuprofen and indomethacin after the first or third courses, whereas ibuprofen was associated with a higher PDA closure rate than indomethacin after the second course (*p* = 0.038; Table S8).

#### 3. Route of Administration (Intravenous or Oral)

For intravenous administration, the PDA closure rate was significantly lower in the second course than in the first course (1st: 63% vs 2nd: 41%; *p* < 0.001; Table S7; Figs. S5 and S6). Multilevel meta-regression analysis similarly showed that a greater number of COX inhibitor courses was associated with lower PDA closure rates (odds ratio, 0.51; 95% CI, 0.44–0.60; *p* < 0.001; Table S6). In contrast, for oral administration, the PDA closure rate did not differ significantly between the first and second courses (1st: 62% vs 2nd: 59%; *p* = 0.762; Tables S6 and S7, Figs. S7 and S8). The PDA closure rate after the third oral course was significantly lower than that after the first course; however, the estimate for the third course was based on only one study (Table S7, Figs. S7 and S8). Substantial heterogeneity was observed in most of these analyses (*I²* = 66.4%–91.2%; Figs. S5 and S7), except after the second oral course (*I²* = 34.9%). Pairwise comparisons between administration routes demonstrated that the PDA closure rate was significantly higher with oral than with intravenous administration after the second course (*p* = 0.0012; Table S8).

### Publication Bias

Funnel plots and Egger’s tests are presented in Figs. S9–11. Egger’s tests indicated significant funnel-plot asymmetry for the third course of COX inhibitors in the overall preterm infant population, based on 10 studies (Fig. S9C).

### Secondary Outcome Measurement: Adverse Events

Our search and screening identified only three studies that reported adverse events after the second or third course of COX inhibitor treatment [14, 19, 20]. These adverse events are summarized in Table S9. Overall, sample sizes were small, and the incidence of adverse events was low. Furthermore, no statistical analyses were performed to compare the risk of adverse events across COX inhibitor courses in these studies.

### Certainty of Evidence (GRADE Assessment)

The overall certainty of evidence for the outcomes is summarized in Table S10. The certainty of evidence for the primary outcome (PDA closure rate) was low. The certainty of evidence for the secondary outcome (adverse events) was very low because of a serious risk of bias.

## DISCUSSION

The evidence from this systematic review suggests that repeated courses of COX inhibitors are associated with lower PDA closure rates than the initial course. The certainty of this evidence was low because of substantial heterogeneity across studies, limited sample sizes, and the low quality of the observational studies. Data on adverse events were very limited, preventing any definitive conclusions. To the best of our knowledge, this study represents the most comprehensive investigation to date of repeated COX inhibitor administration in preterm infants with hemodynamically significant PDA.

Our study revealed that repeated administration of COX inhibitors was associated with a lower PDA closure rate in preterm infants. Possible explanations for this finding include confounding by indication—whereby infants with more severe or refractory PDA were more likely to receive repeated courses—and differences in treatment timing, specifically a higher postnatal age at subsequent courses. Because data on PDA severity and the timing of drug administration were limited, further analyses adjusting for these factors were not feasible. Although the forest plots showed no apparent temporal changes in PDA closure rates from 2002 to 2026, the increasing adoption of expectant management warrants continued monitoring of closure rates as clinical practice evolves [3, 4].

Our subgroup analyses showed that EPT/ELBW infants had lower PDA closure rates than preterm infants from non-overlapping cohorts (Table S8). This finding aligns with previous clinical observations; for example, a study reported lower PDA closure rates after ibuprofen treatment among infants born at 23–26 weeks’ gestation compared with those born at 27–30 weeks’ gestation [45]. These results highlight that extreme prematurity remains a major clinical challenge for pharmacological PDA closure.

Our subgroup analysis demonstrated similar PDA closure rates between ibuprofen and indomethacin in the first and third courses, whereas ibuprofen was associated with a higher closure rate in the second course (Table S8). Given that both agents have generally shown similar PDA closure rates [46, 47], this difference may reflect confounding by indication. The more favorable adverse-effect profile of ibuprofen [46] may have led clinicians to preferentially use it in infants perceived to have a lower risk of treatment failure.

Unlike other subgroups, oral administration showed no significant decline in PDA closure rates after the second course compared with the first (Table S7; Fig. S8). This finding may partly reflect developmental changes in gastrointestinal drug absorption with increasing postnatal age [48], which could influence ibuprofen exposure and contribute to the relatively preserved closure rates after the second course. The higher PDA closure rate with oral than with intravenous administration after the second course (Table S8) may also reflect confounding by indication, such as the preferential use of oral agents in infants with less severe illness.

We observed substantial heterogeneity in outcomes across included studies. This heterogeneity persisted across subgroup analyses, suggesting that factors beyond EPT/ELBW status, drug type, and route of administration may have contributed to the observed variability. This systematic review did not restrict study eligibility by publication year or country. The included studies were published between 2002 and 2026 and represented 15 countries. This geographic and temporal diversity may enhance the generalizability of our findings but may also contribute to between-study heterogeneity. To account for this variability, we employed multilevel mixed-effects meta-regression and within-study pairwise comparisons. Although the pairwise comparisons had reduced statistical power due to the inherent pairwise deletion of studies lacking third-course data, the concurrent use of multilevel meta-regression maximized the use of all available data to robustly evaluate the overall trend.

This systematic review has several limitations. First, our definition of PDA closure encompassed varying criteria across the included studies; therefore, the pooled closure rates primarily reflect clinical resolution without the need for further treatment rather than strict anatomical closure. Accordingly, the definition included both echocardiographically confirmed complete closure and residual PDA without hemodynamic significance that did not require additional treatment. This point should be considered when interpreting the pooled closure rates and applying our findings to treatment decisions for individual patients. Second, our primary focus was to report the observed PDA closure rates relevant to clinical practice, rather than established causal inference. Limited reporting of important covariates, such as treatment timing, precluded analyses that could adequately account for potential confounding and assess the independent effects of repeated COX inhibitor courses on PDA closure. Third, our search strategy may have overlooked studies reporting adverse events. Because eligibility required studies to report PDA closure rates after two or more courses of COX inhibitor treatment, studies reporting adverse events without corresponding closure rates may have been excluded during screening. Fourth, the possibility of publication bias cannot be ruled out. Egger’s test detected statistically significant funnel plot asymmetry for the third course of COX inhibitors (Fig. S9C). However, the limited number of included studies makes it difficult to confirm or exclude publication bias [49]. Despite these limitations, to our knowledge, this is the first systematic review to evaluate PDA closure rates following repeated courses of COX inhibitor treatment in preterm infants. By synthesizing evidence across clinically relevant subgroups, including extremely preterm/extremely low birth weight infants, drug type, and route of administration, our findings provide evidence to inform clinical decision-making.

## CONCLUSION

This systematic review and meta-analysis showed that PDA closure rates were lower after repeated courses of COX inhibitors than after the initial course in preterm infants, except for the second course of oral administration. Data on the safety of repeated courses were limited.

## Data Availability

All data produced in the present work are contained in the manuscript.

## Acknowledgments

We thank members of the Japan Evidence-Based Neonatology Research Group for their support.

## Author Contributions

J.S. conceived and designed the study. J.S. and K.O. developed the search strategy. J.S. and YS.W. performed the title and abstract screening and full-text screening. J.S. and K.Y. performed the analyses. J.S., YS.W., K.Y., N.K., T.I, and K.T. interpreted the data and drafted the manuscript. All authors reviewed the manuscript and approved the final manuscript.

## Statement of Ethics

As this systematic review was based solely on published literature, patient consent was not required.

## Data Availability Statement

All data associated with this study are included in the paper.

## List of Supplementary Figures and Legends

**Figure S1.**
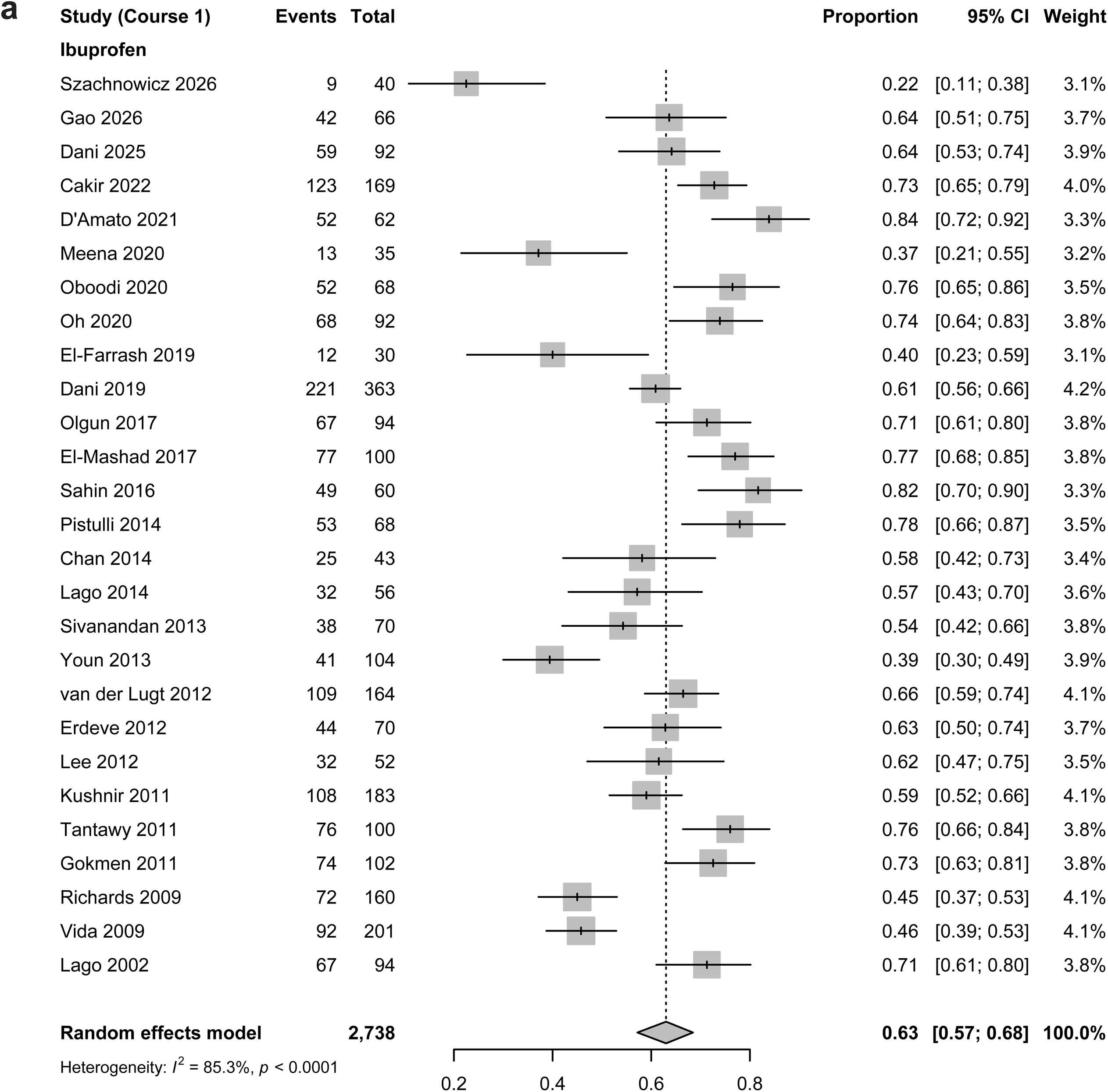
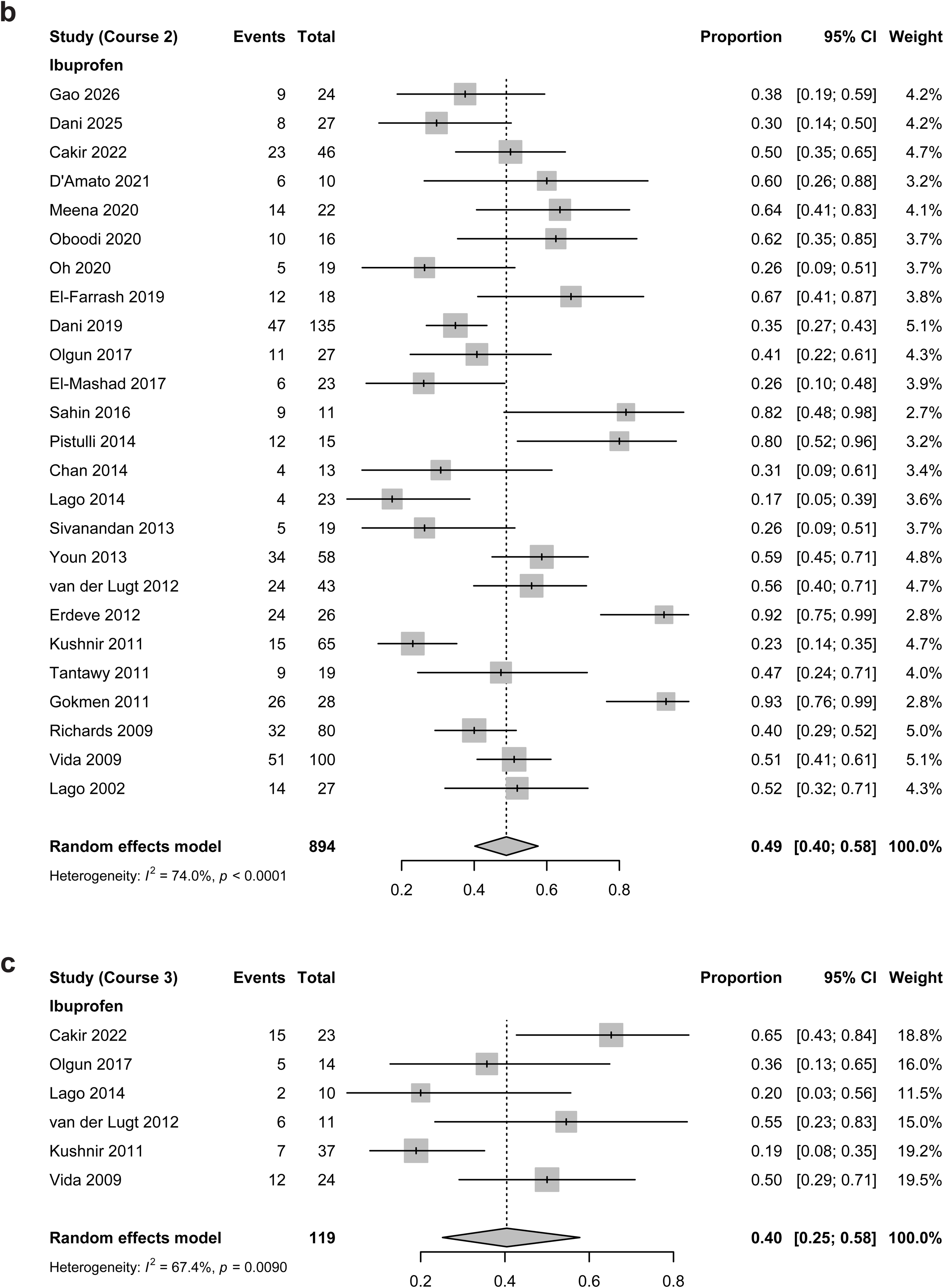
Forest plots of PDA closure rates after ibuprofen treatment. **a.** Closure rate after the first course. **b.** Closure rate after the second course. **c.** Closure rate after the third course.

**Figure S2.**
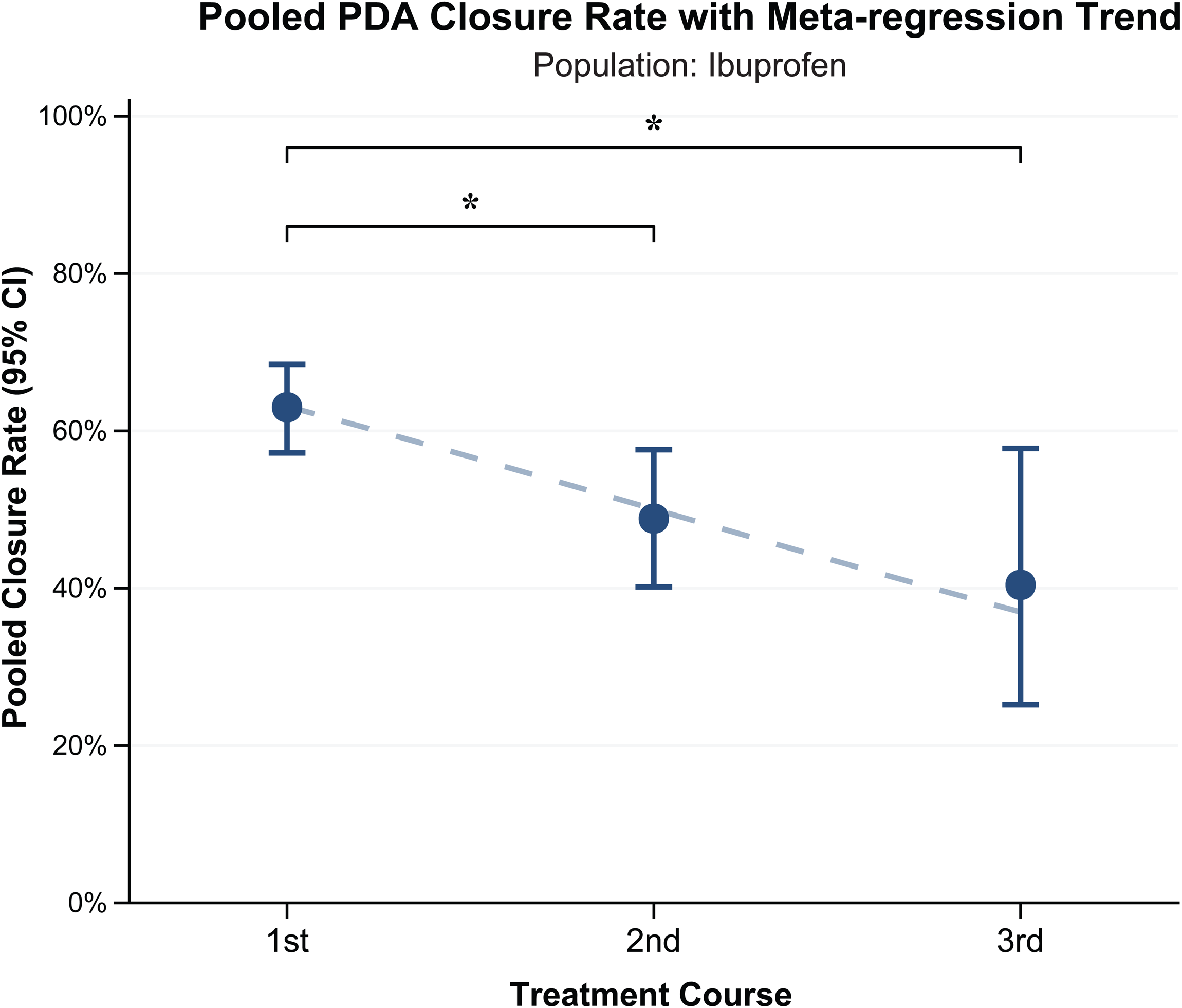
Pooled PDA closure rates and meta-regression trends after ibuprofen treatment. Pooled PDA closure rates are shown after the first, second, and third treatment courses, with corresponding 95% confidence intervals (CIs). Meta-regression analysis illustrates temporal trends in PDA closure rates across treatment courses.

**Figure S3.**
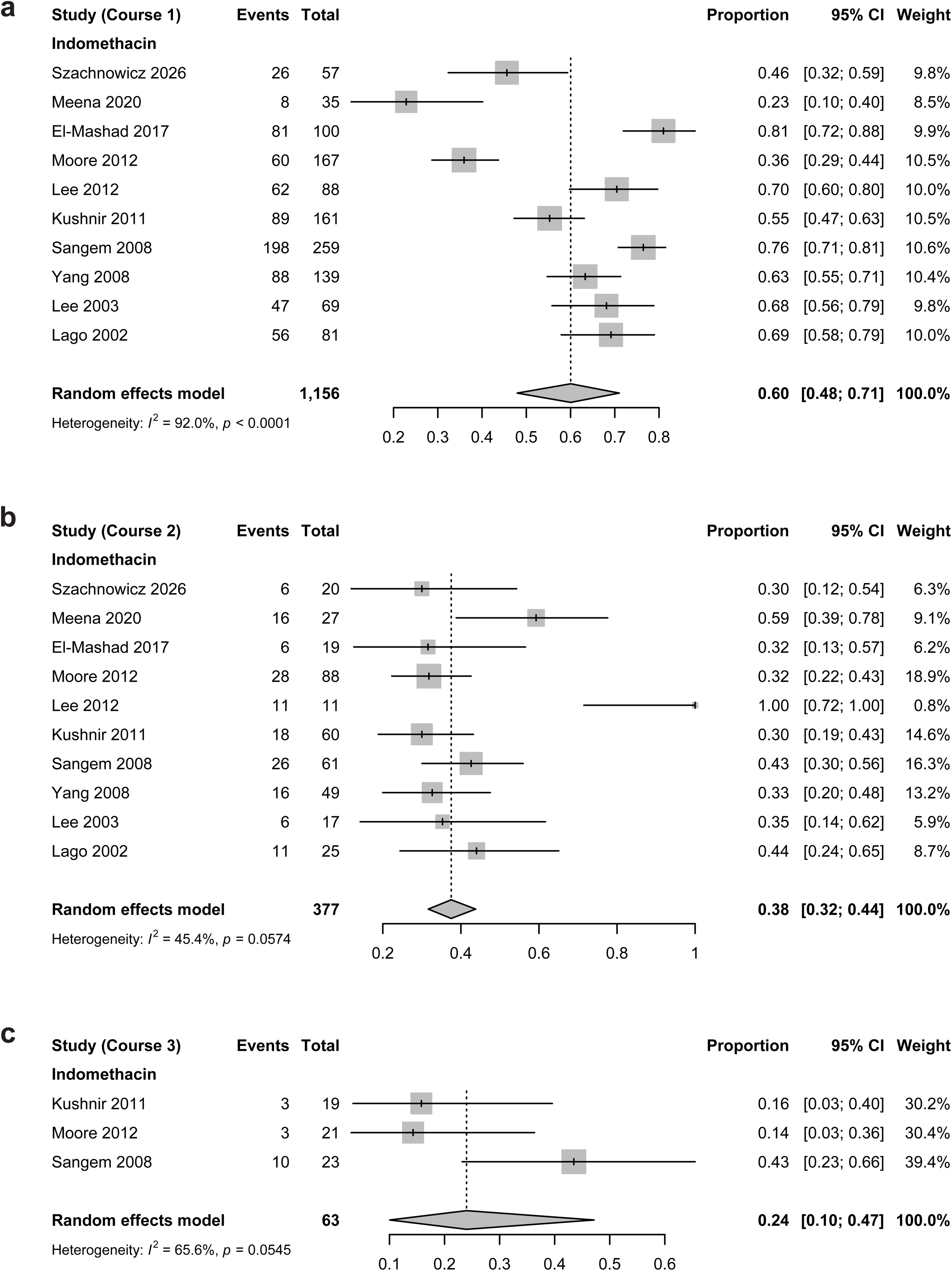
Forest plots of PDA closure rates after indomethacin treatment. **a.** Closure rate after the first course. **b.** Closure rate after the second course. **c.** Closure rate after the third course.

**Figure S4.**
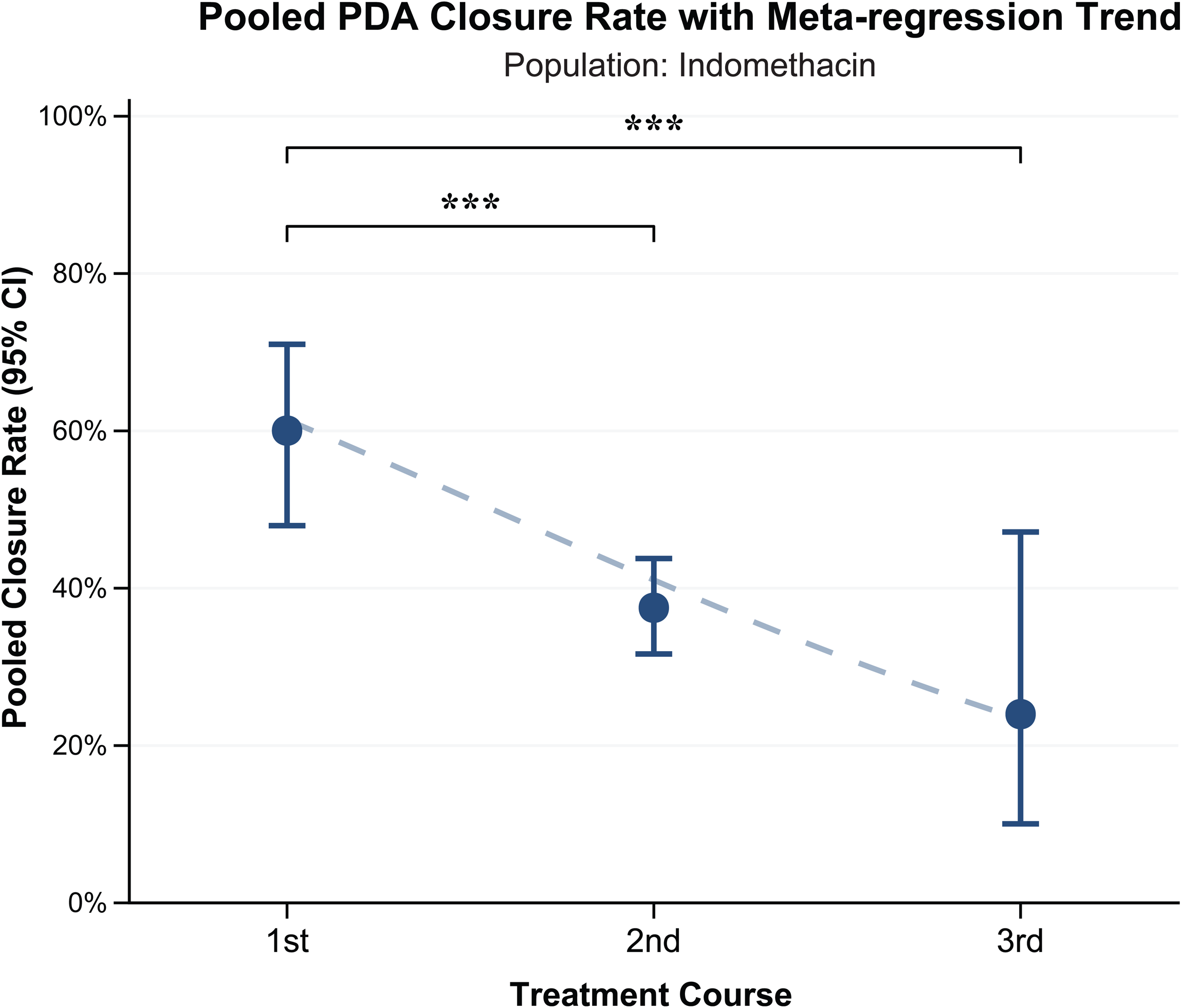
Pooled PDA closure rates and meta-regression trends after indomethacin treatment. Pooled PDA closure rates are shown after the first, second, and third treatment courses, with corresponding 95% confidence intervals (CIs). Meta-regression analysis illustrates temporal trend in PDA closure rates across treatment courses.

**Figure S5.**
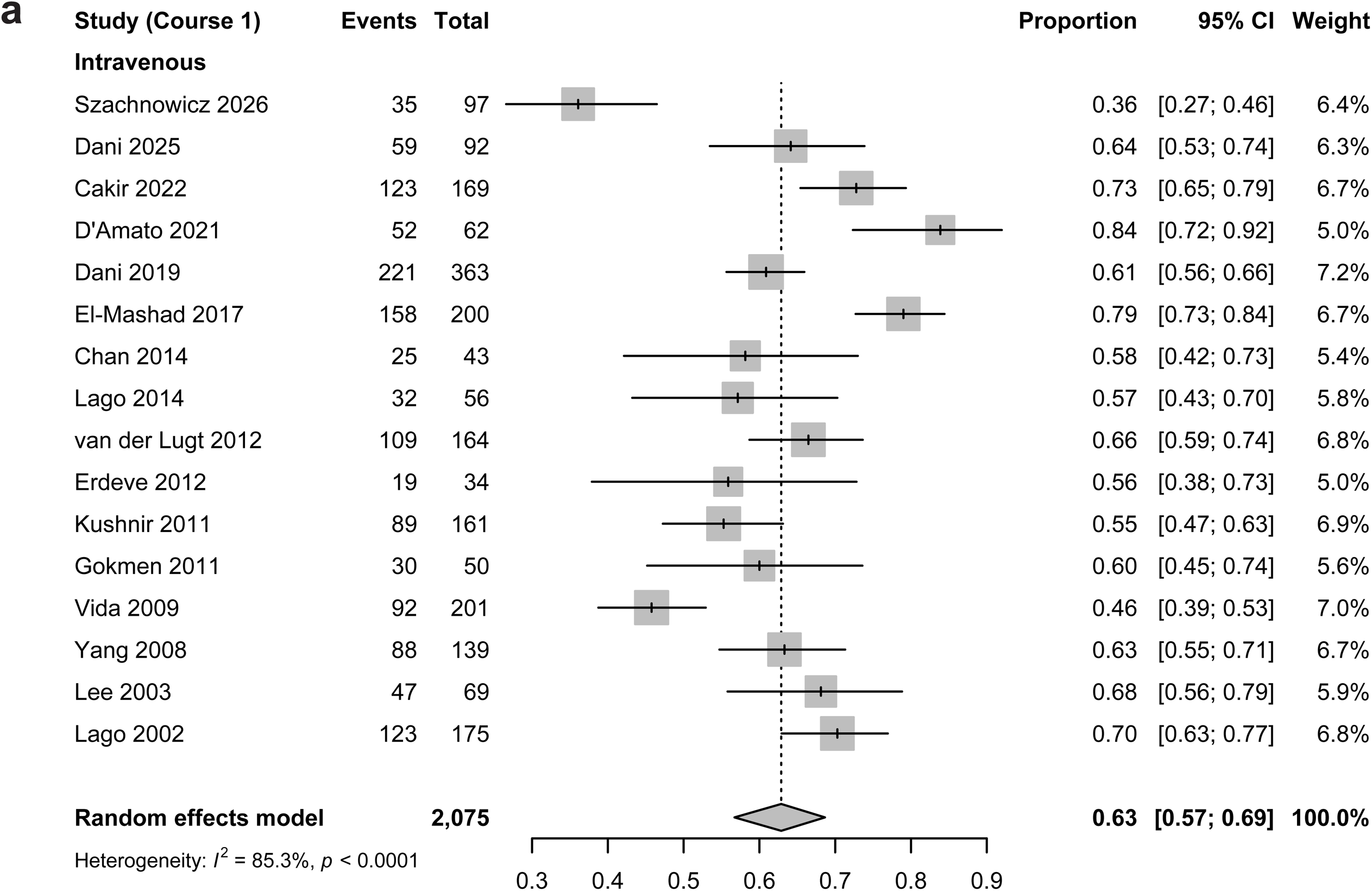
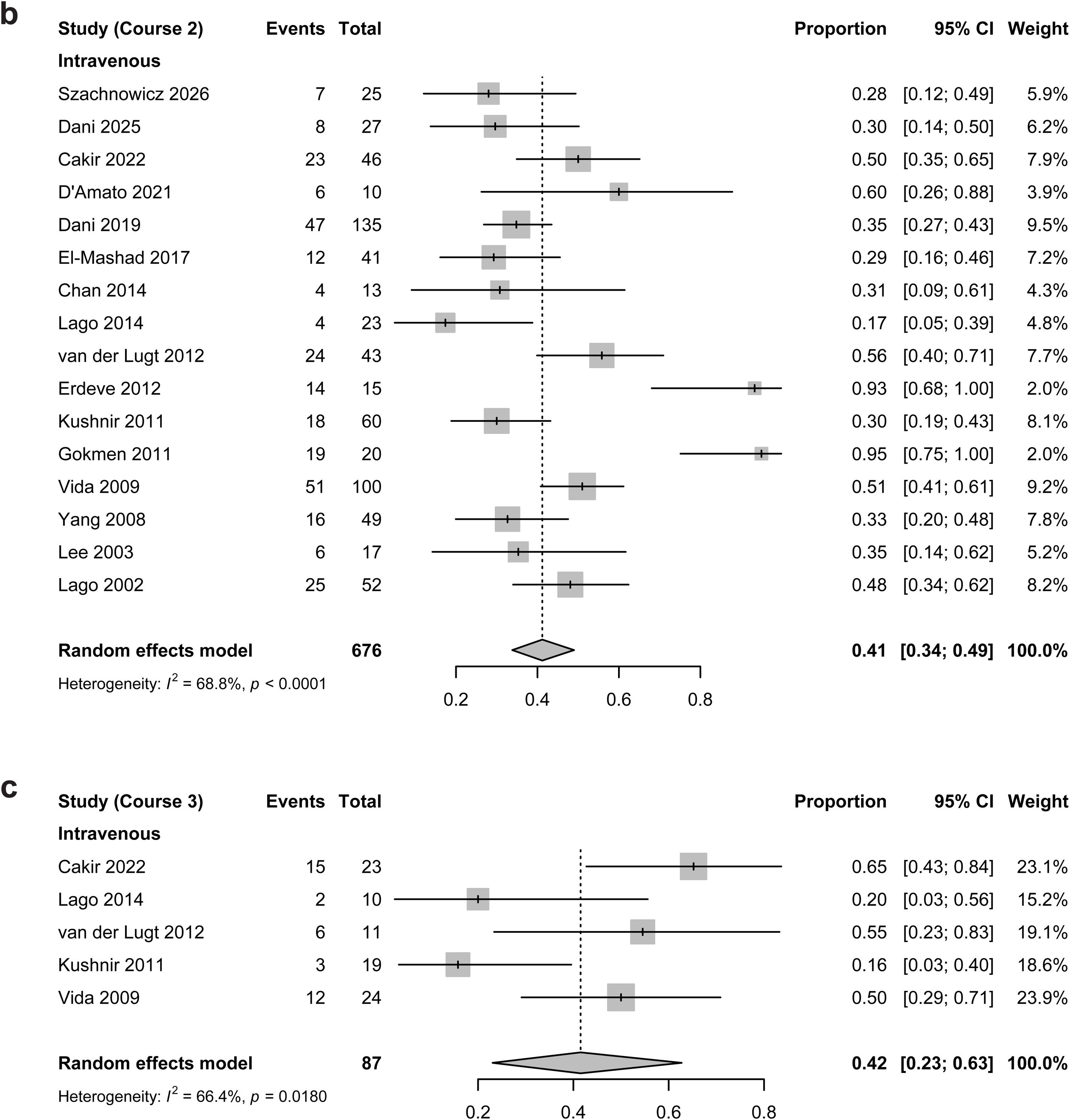
Forest plots of PDA closure rates after intravenous COX inhibitor administration. **a.** Closure rate after the first course. **b.** Closure rate after the second course. **c.** Closure rate after the third course.

**Figure S6.**
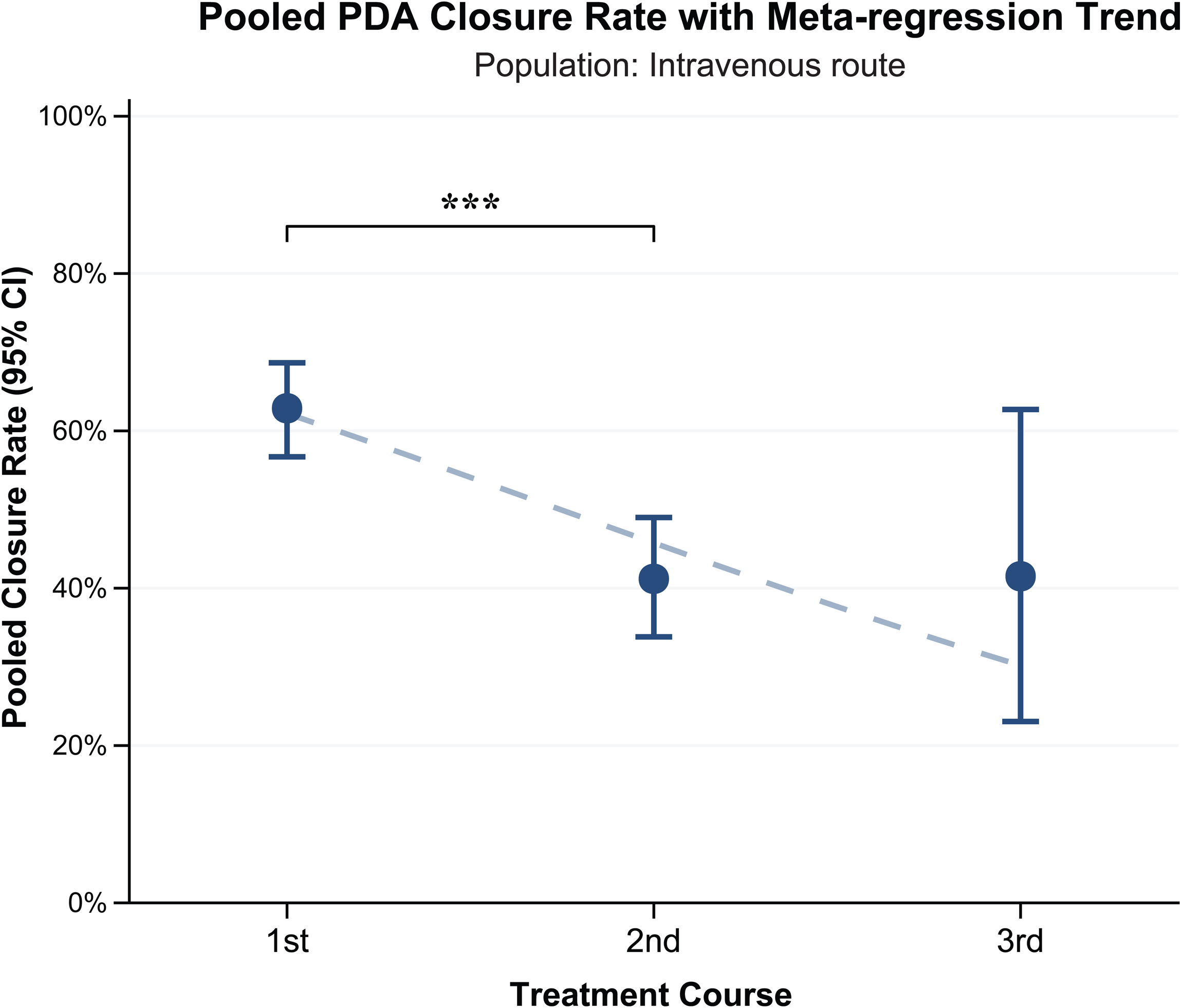
Pooled PDA closure rates and meta-regression trends after intravenous COX inhibitor administration. Pooled PDA closure rates are shown after the first, second, and third treatment courses, with corresponding 95% confidence intervals (CIs). Meta-regression analysis illustrates temporal trend in PDA closure rates across treatment courses.

**Figure S7.**
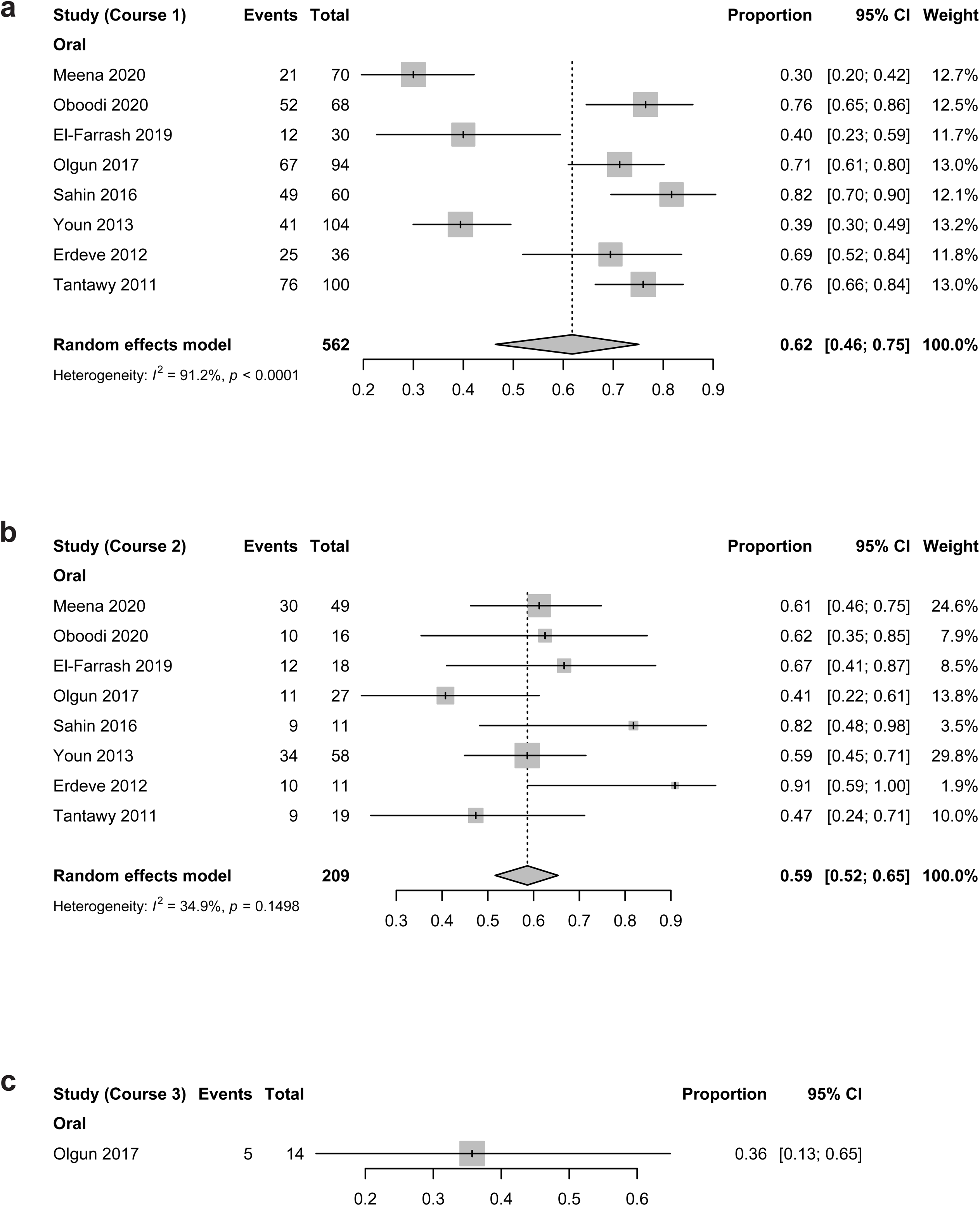
Forest plots of PDA closure rates after oral COX inhibitor administration. **a.** Closure rate after the first course. **b.** Closure rate after the second course. **c.** Closure rate after the third course.

**Figure S8.**
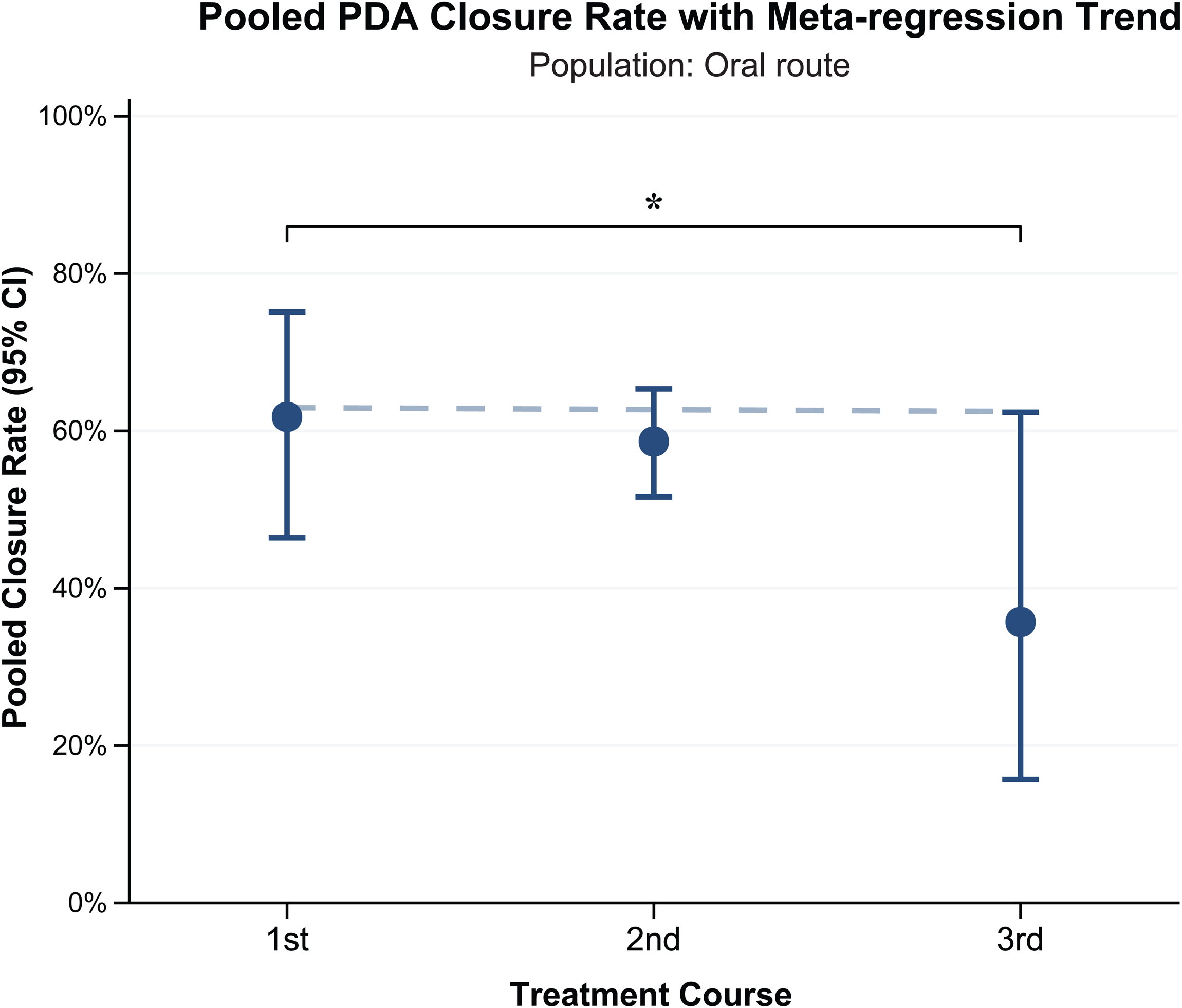
Pooled PDA closure rates and meta-regression trends after oral COX inhibitor administration. Pooled PDA closure rates are shown after the first, second, and third treatment courses, with corresponding 95% confidence intervals (CIs). Meta-regression analysis illustrates temporal trend in PDA closure rates across treatment courses.

**Figure S9.**
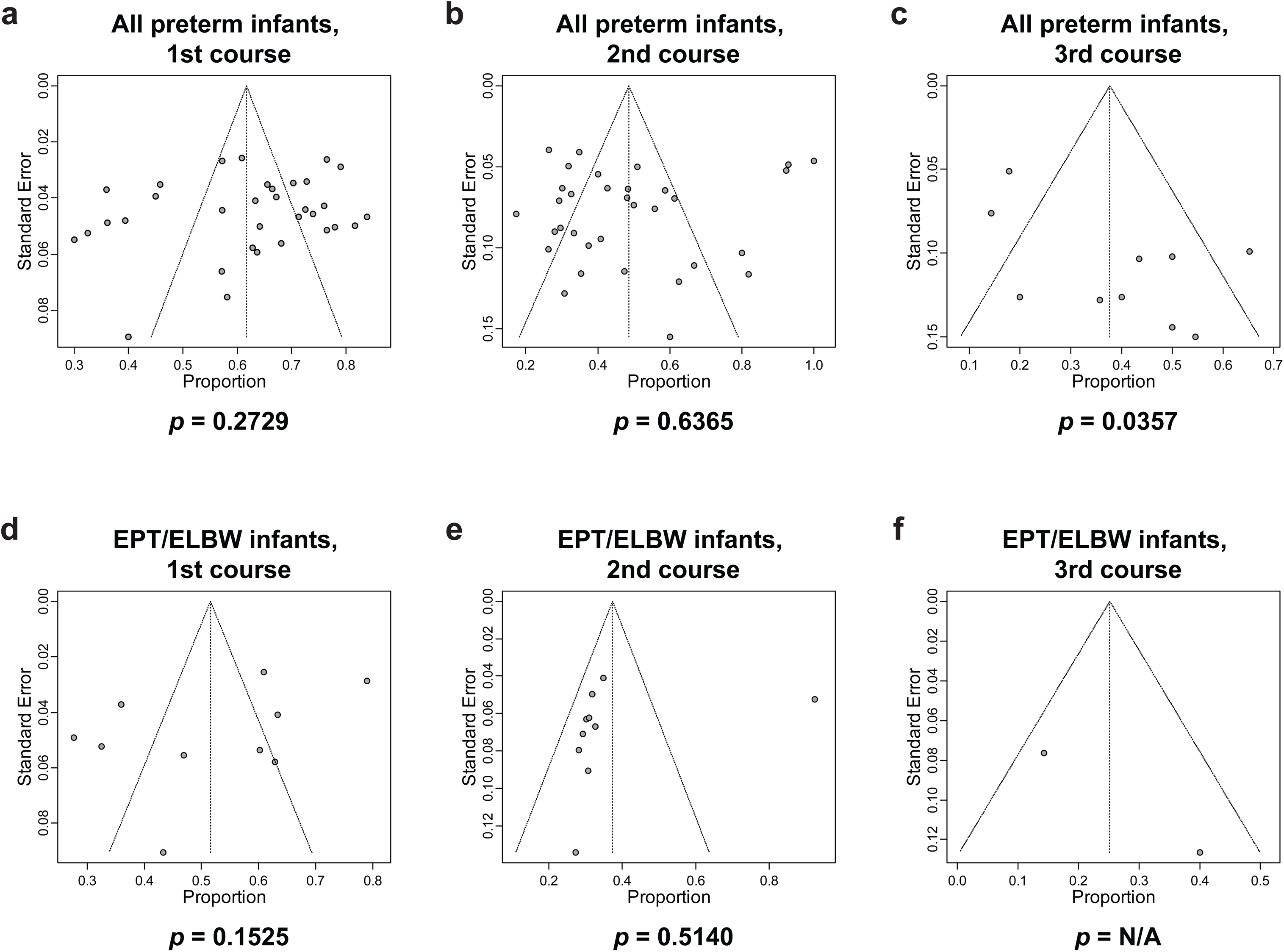
Funnel plots and Egger’s tests for publication bias in the analysis of PDA closure rates after COX inhibitor treatment. a–c. Overall preterm infant population after the first, second, and third courses, respectively. **d–f.** EPT/ELBW infants after the first, second, and third courses, respectively.

**Figure S10.**
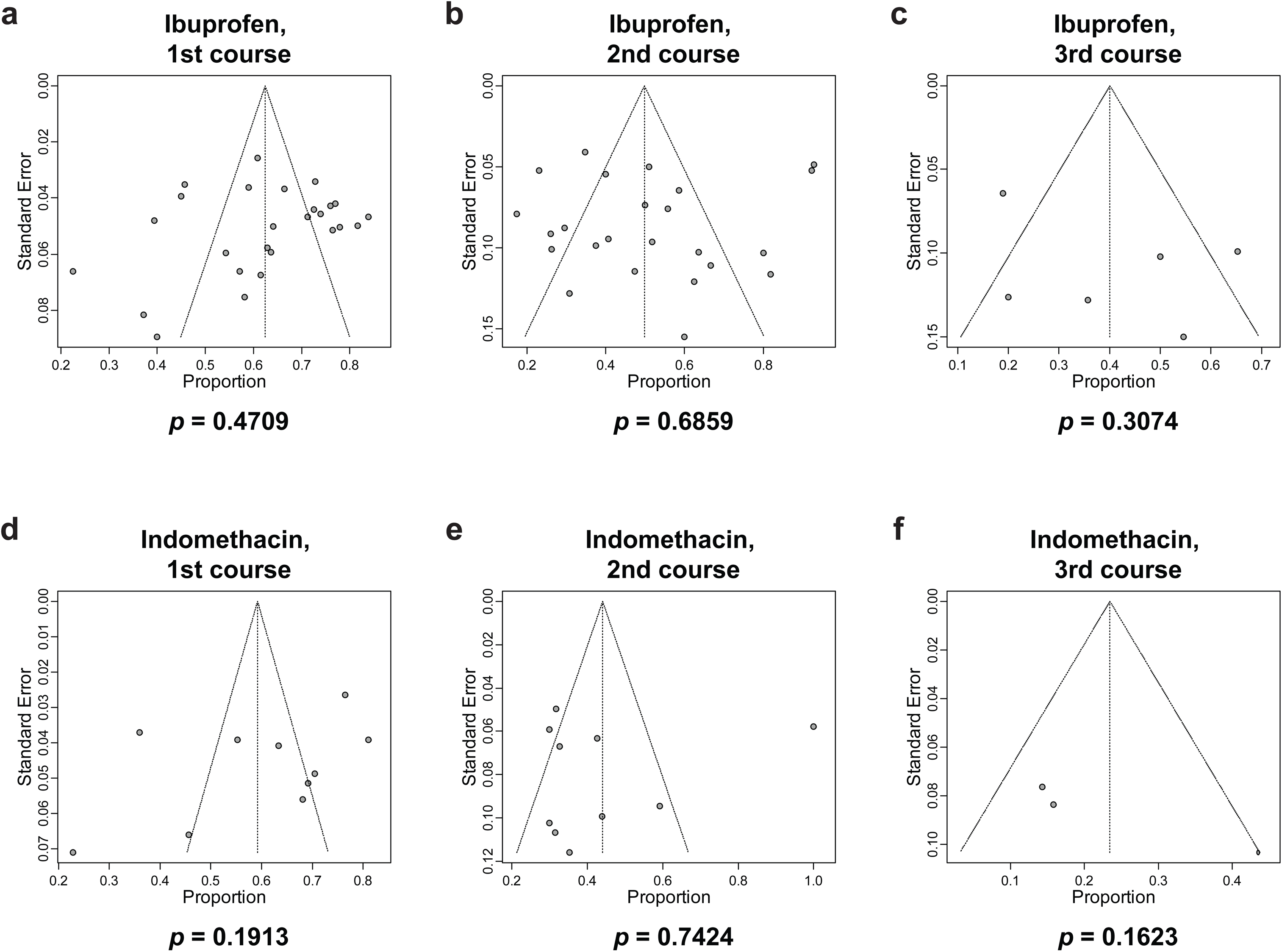
Funnel plots and Egger’s tests for publication bias in the analysis of PDA closure rates after COX inhibitor treatment. a–c. Ibuprofen-treated infants after the first, second, and third courses, respectively. **d–f.** Indomethacin-treated infants after the first, second, and third courses, respectively.

**Figure S11.**
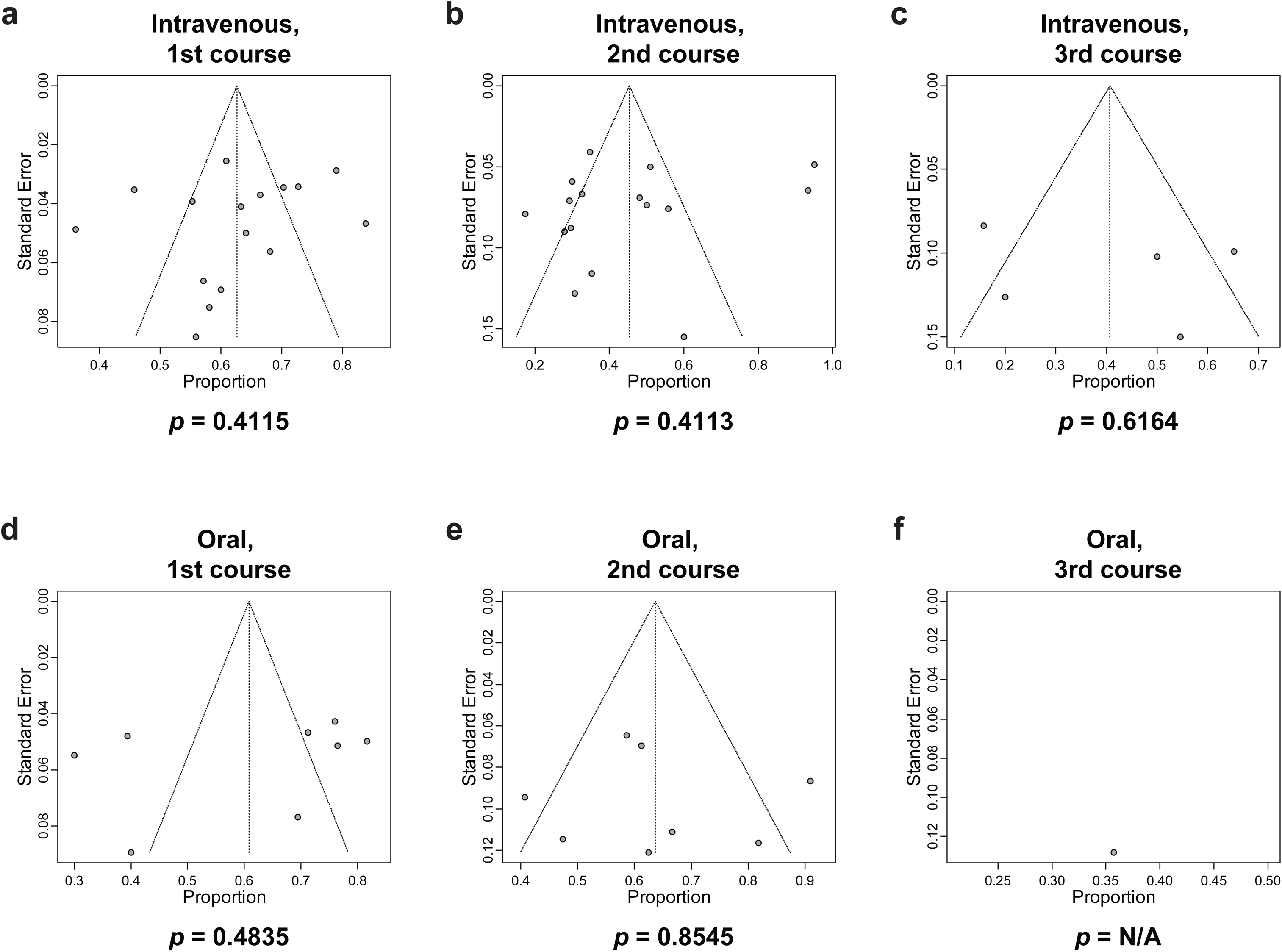
Funnel plots and Egger’s tests for publication bias in the analysis of PDA closure rates after COX inhibitor treatment. a–c. Infants receiving intravenous administration after the first, second, and third courses, respectively. **d–f.** Infants receiving oral administration after the first, second, and third courses, respectively.

## List of Supplementary Tables

**Table S1.**
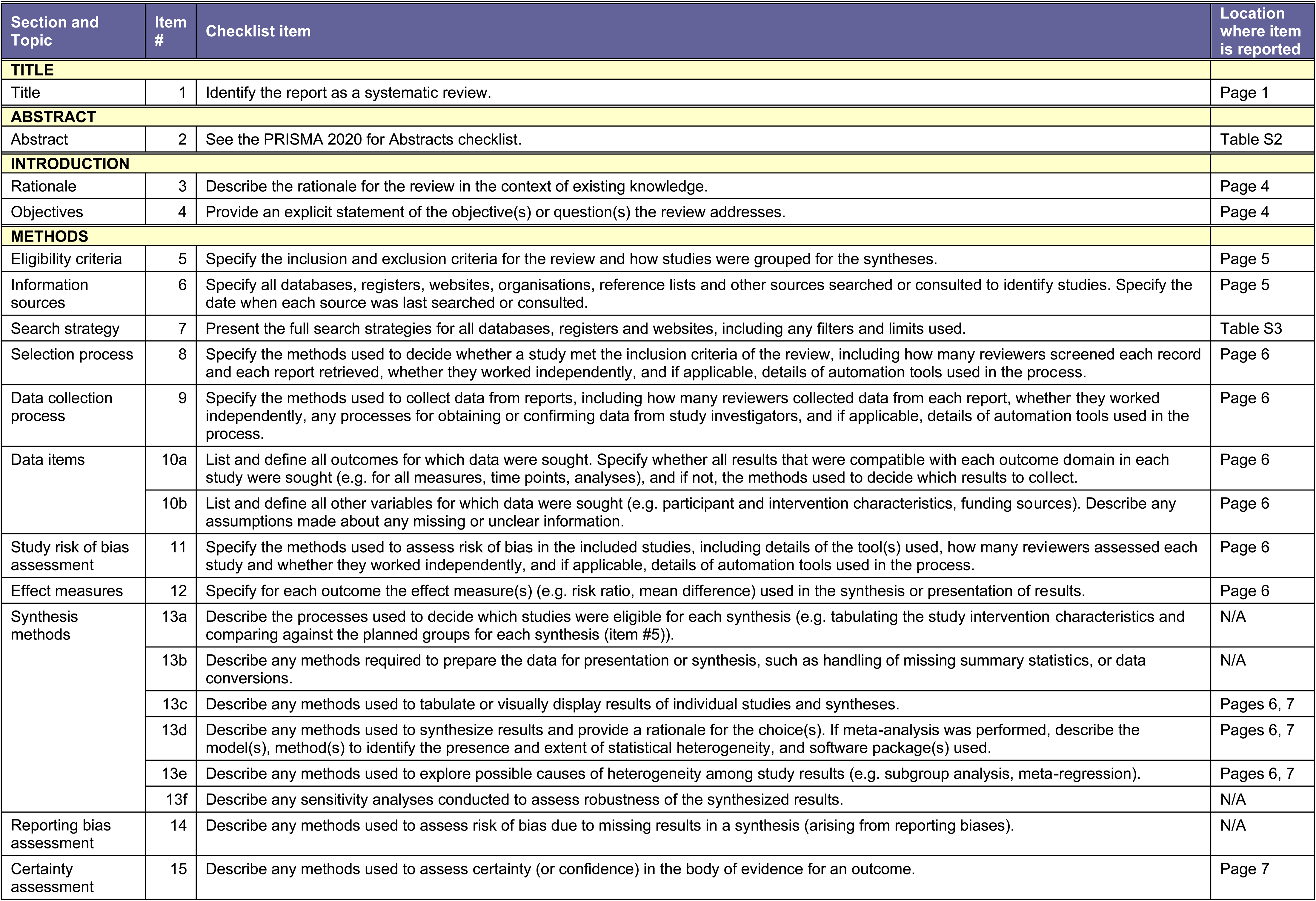
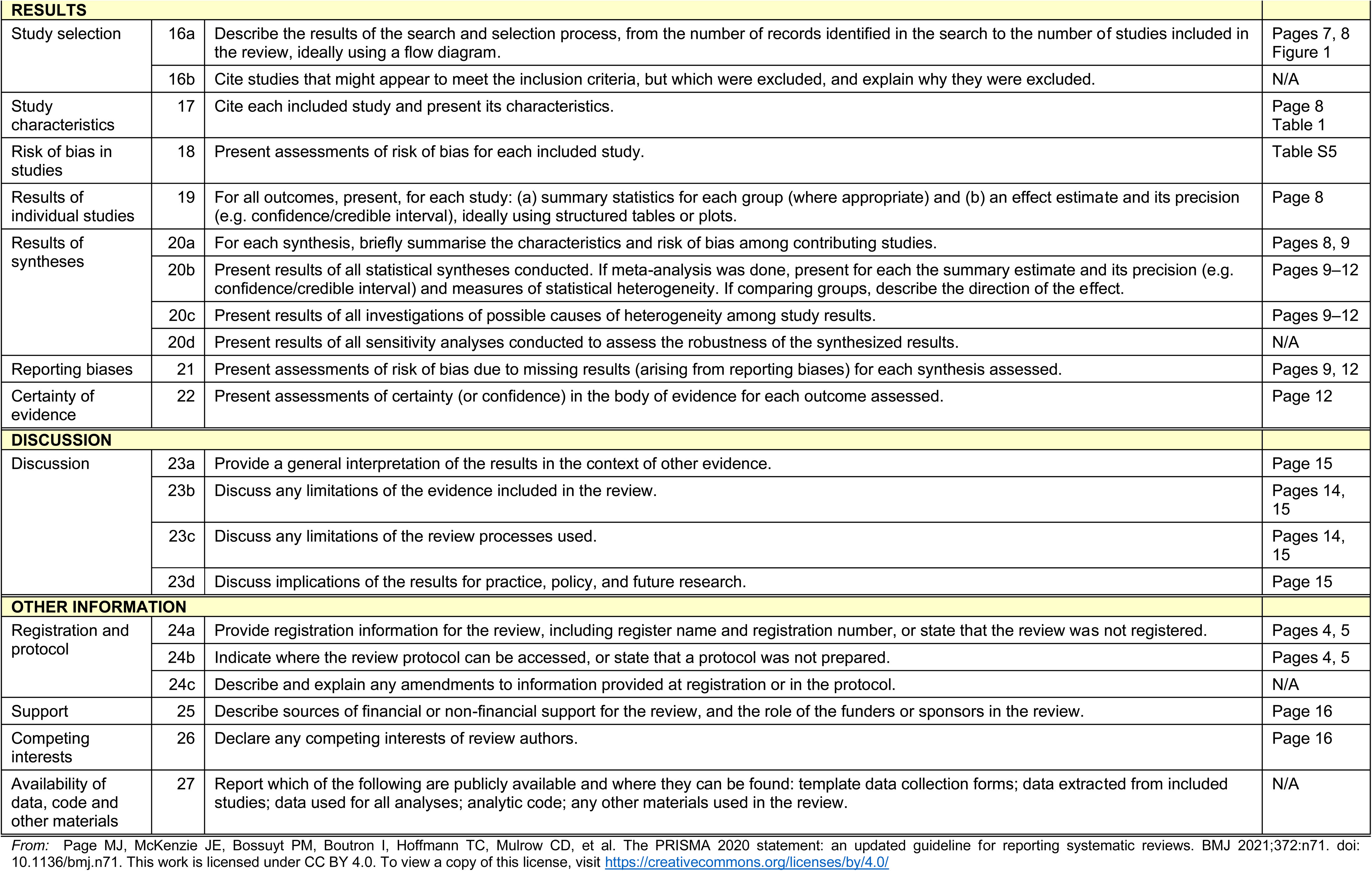
PRISMA 2020 Checklist.

**Table S2.** PRISMA 2020 for Abstracts Checklist.

| Section and Topic | Item # | Checklist item | Reported (Yes/No) |
| --- | --- | --- | --- |
| **TITLE** |  |  |  |
| Title | 1 | Identify the report as a systematic review. | Yes |
| **BACKGROUND** |  |  |  |
| Objectives | 2 | Provide an explicit statement of the main objective(s) or question(s) the review addresses. | Yes |
| **METHODS** |  |  |  |
| Eligibility criteria | 3 | Specify the inclusion and exclusion criteria for the review. | Yes |
| Information sources | 4 | Specify the information sources (e.g. databases, registers) used to identify studies and the date when each was last searched. | Yes |
| Risk of bias | 5 | Specify the methods used to assess risk of bias in the included studies. | Yes |
| Synthesis of results | 6 | Specify the methods used to present and synthesise results. | Yes |
| **RESULTS** |  |  |  |
| Included studies | 7 | Give the total number of included studies and participants and summarise relevant characteristics of studies. | Yes |
| Synthesis of results | 8 | Present results for main outcomes, preferably indicating the number of included studies and participants for each. If meta-analysis was done, report the summary estimate and confidence/credible interval. If comparing groups, indicate the direction of the effect (i.e. which group is favoured). | Yes |
| **DISCUSSION** |  |  |  |
| Limitations of evidence | 9 | Provide a brief summary of the limitations of the evidence included in the review (e.g. study risk of bias, inconsistency and imprecision). | Yes |
| Interpretation | 10 | Provide a general interpretation of the results and important implications. | Yes |
| **OTHER** |  |  |  |
| Funding | 11 | Specify the primary source of funding for the review. | N/A |
| Registration | 12 | Provide the register name and registration number. | Yes |

From: Page MJ, McKenzie JE, Bossuyt PM, Boutron I, Hoffmann TC, Mulrow CD, et al. The PRISMA 2020 statement: an updated guideline for reporting systematic reviews. BMJ 2021;372:n71. doi: 10.1136/bmj.n71. This work is licensed under CC BY 4.0. To view a copy of this license, visit https://creativecommons.org/licenses/by/4.0/

**Table S3.**
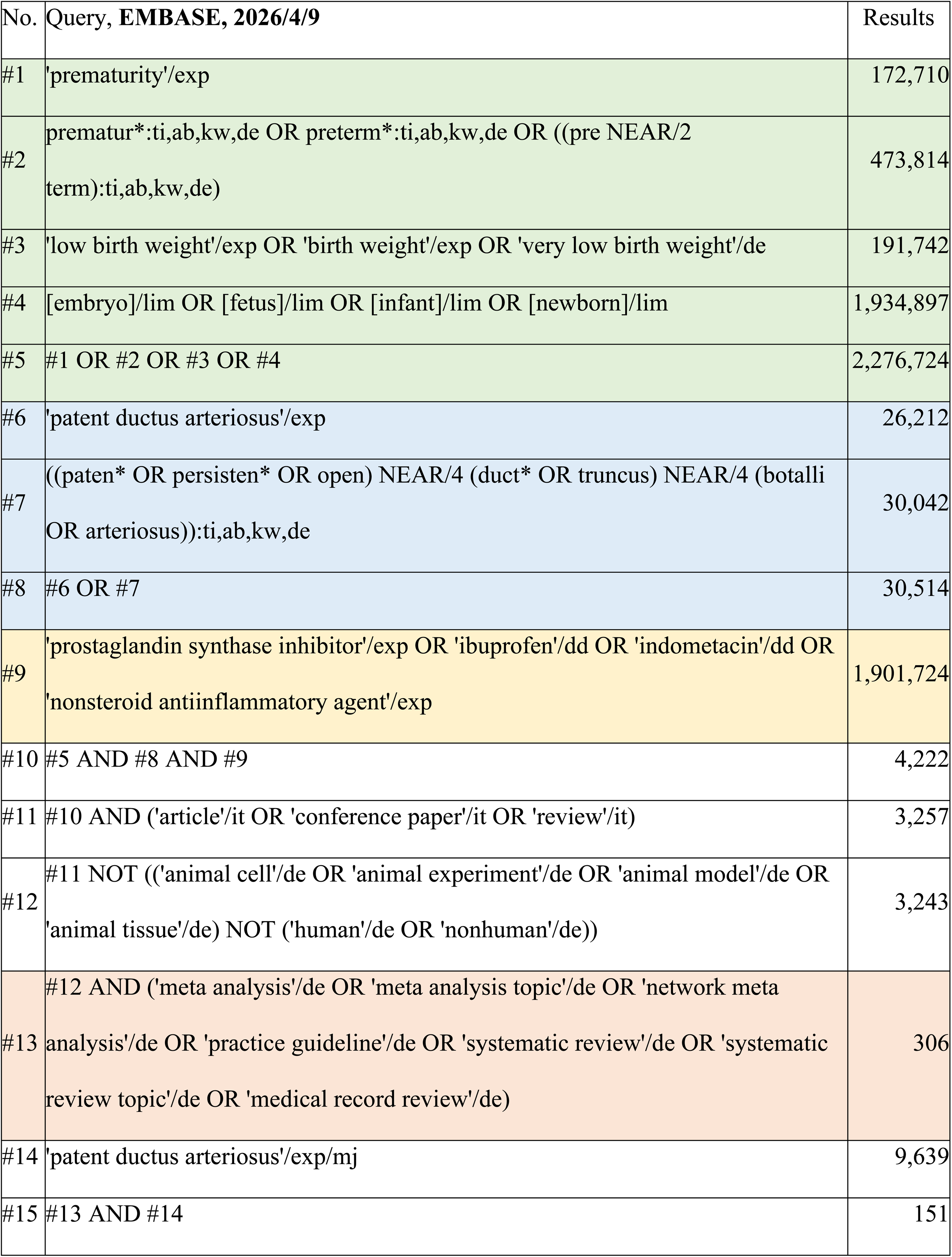
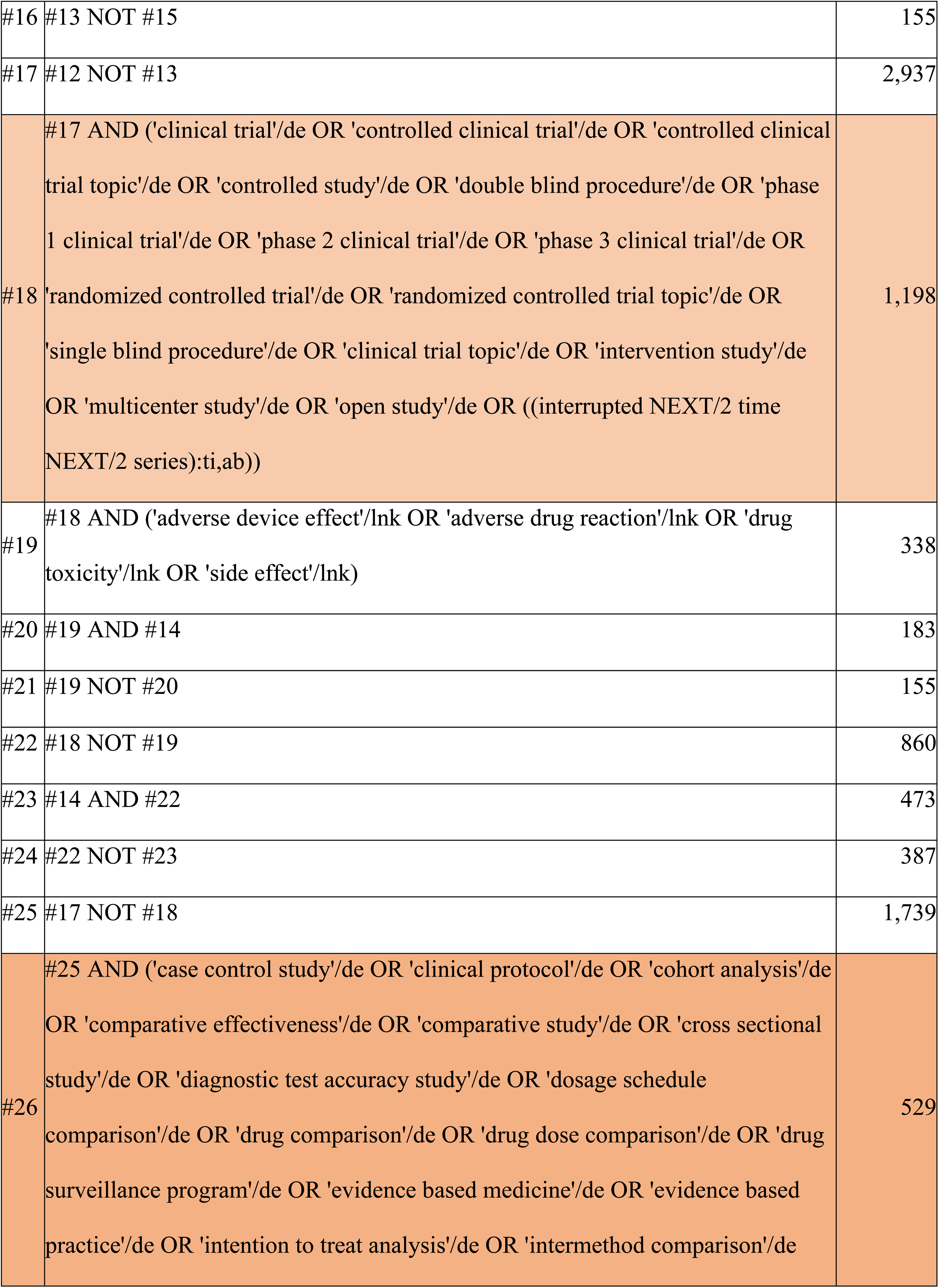
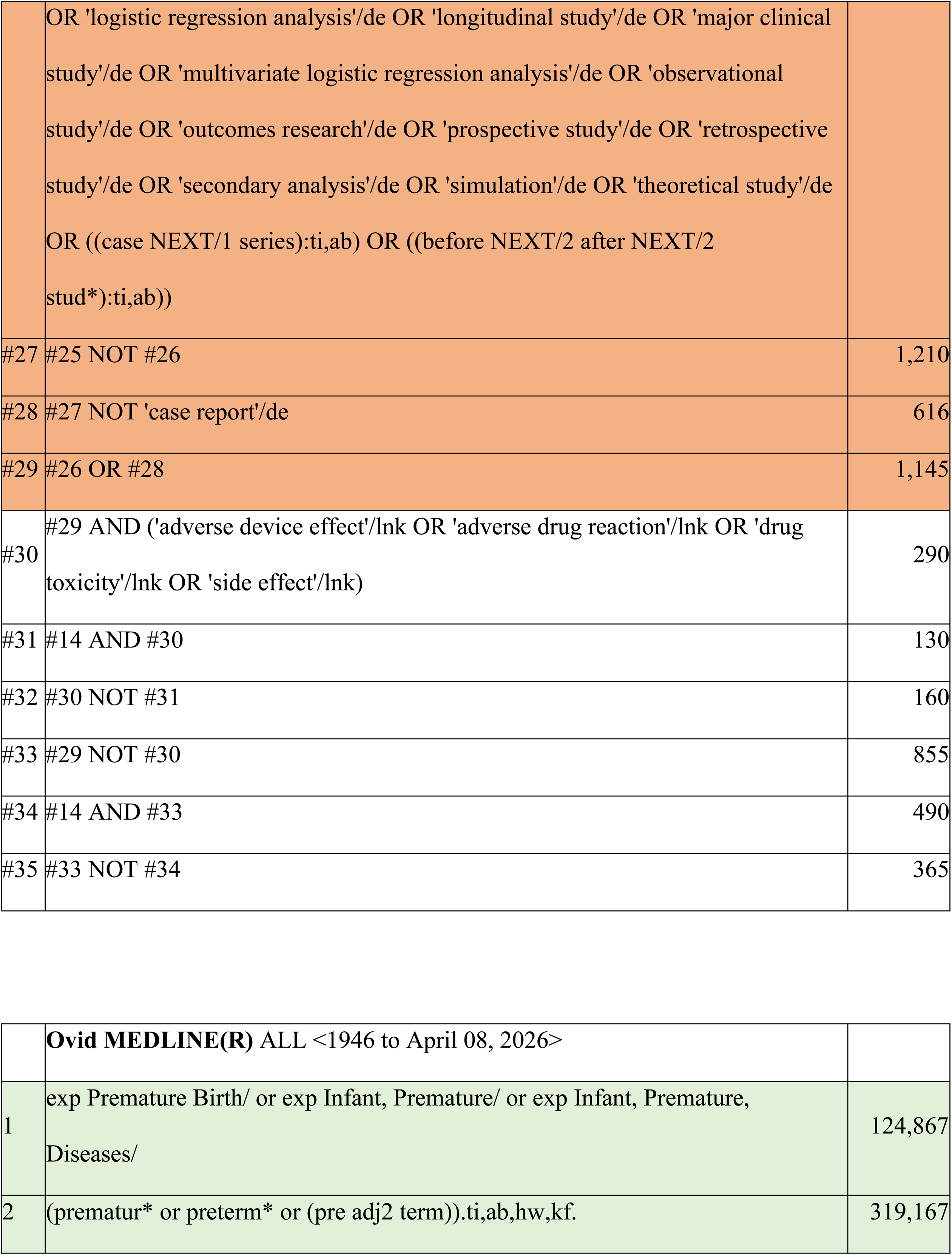
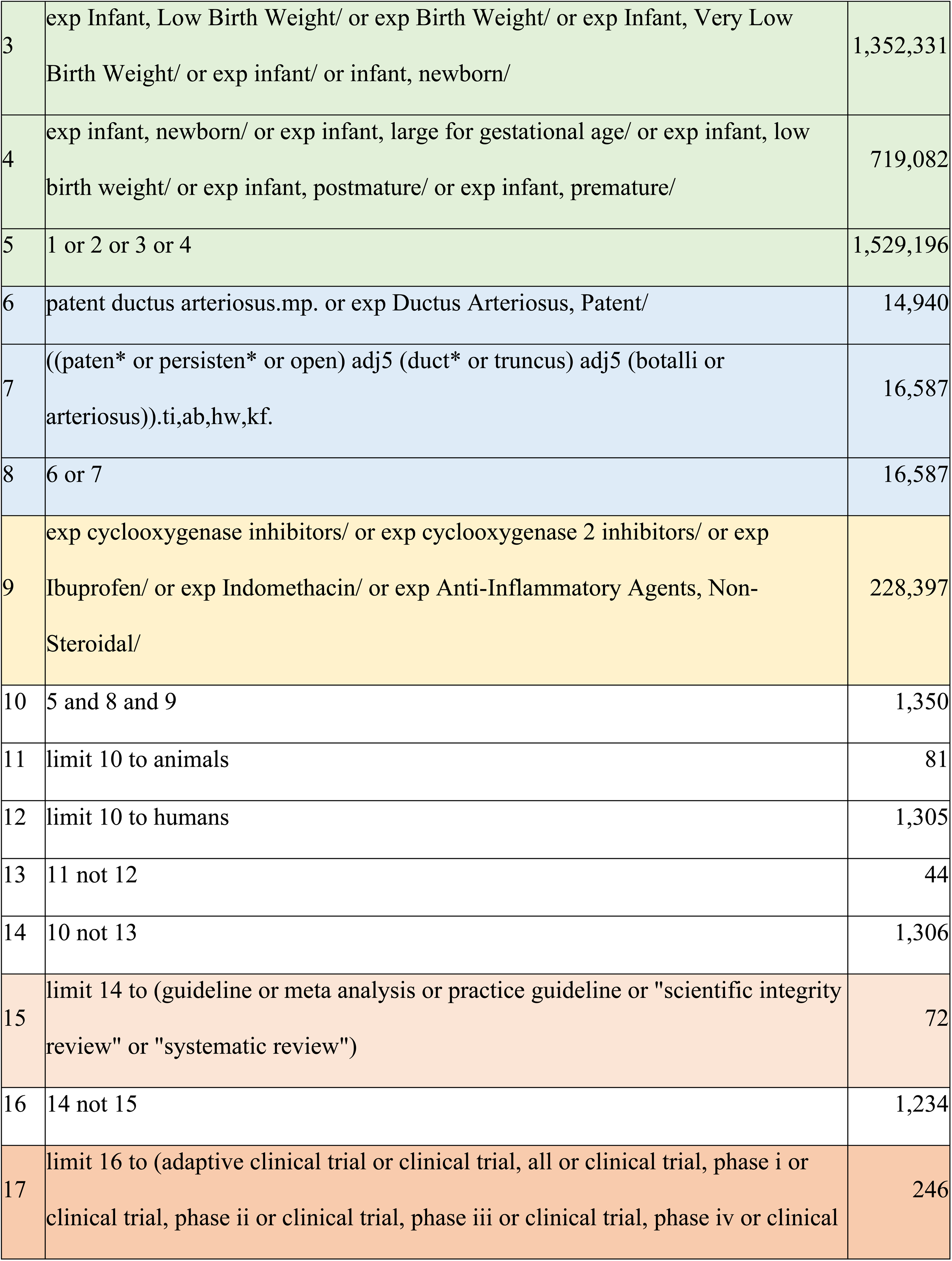
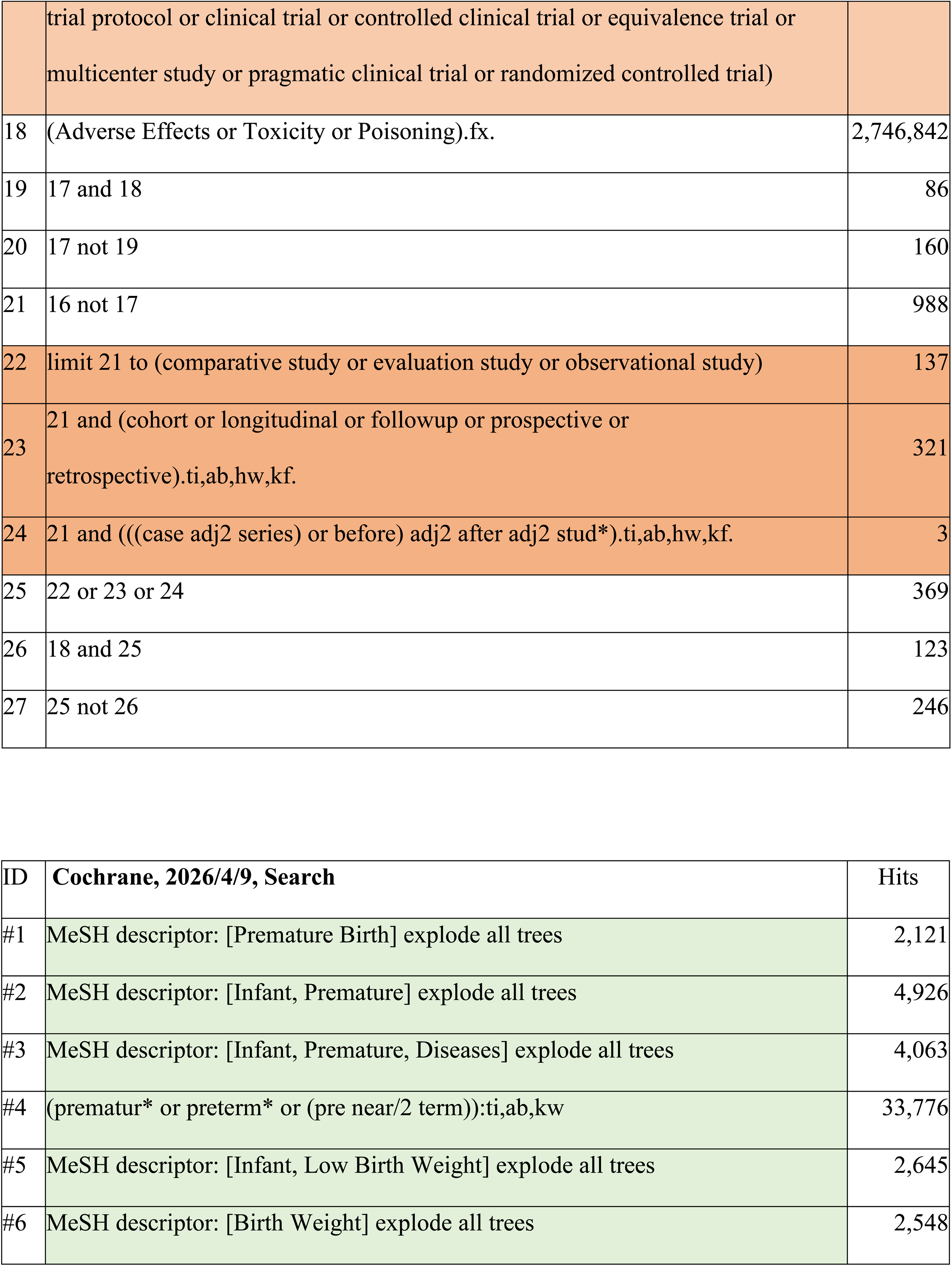
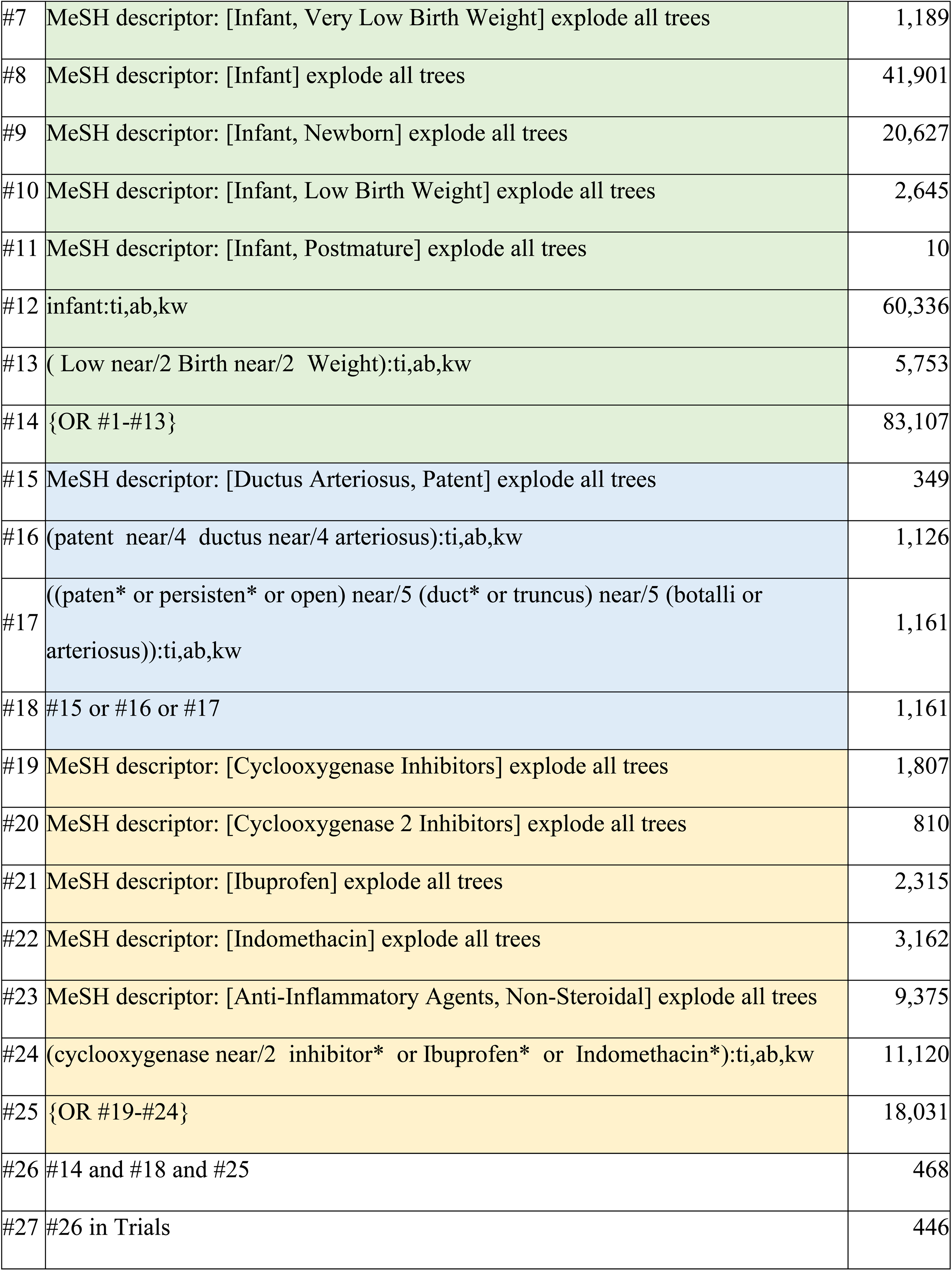
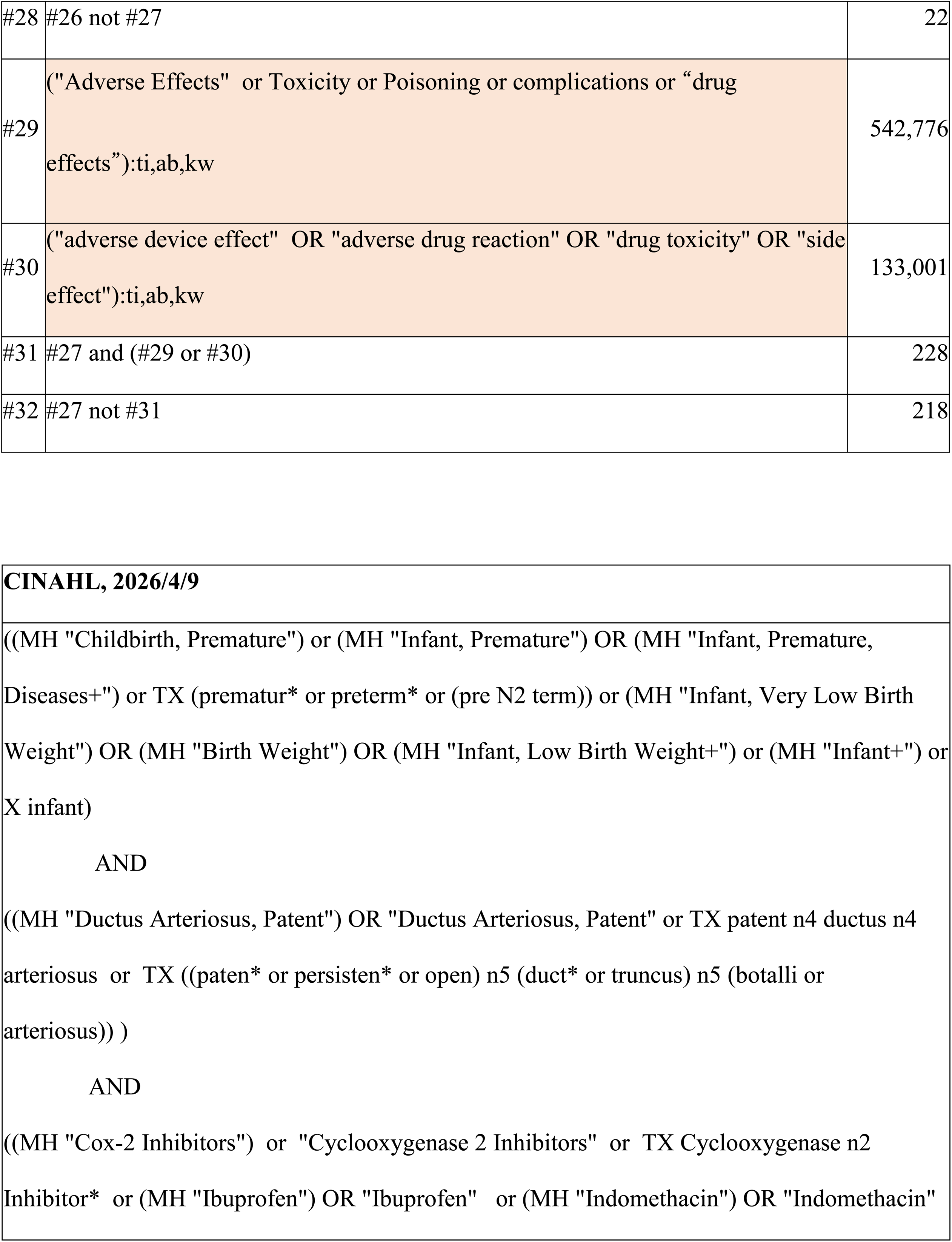
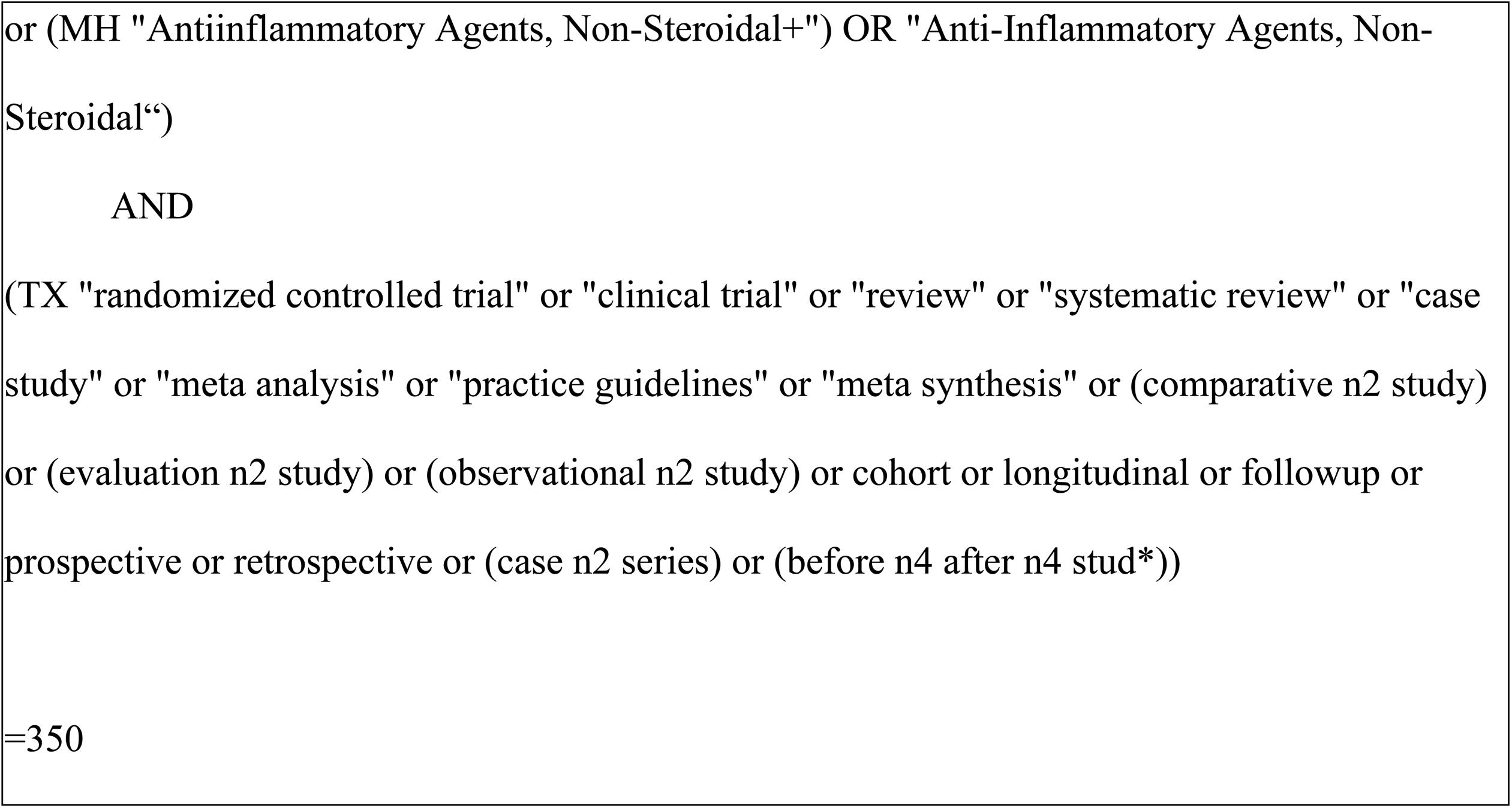
Search terms used in each database.

**Table S4.** Detailed characteristics of the included studies.

| Author,<br>Publication Year | Study Period | Country | Inclusion Criteria | Drug | Route | Cohort size,<br>number | Male,<br>number | GA, weeks | Birth weight, g | 1st course of<br>COX inhibitors,<br>number | PDA closure<br>after the 1st course,<br>number | 2nd course of<br>COX inhibitors,<br>number | PDA closure<br>after the 2nd course,<br>number | 3rd course of<br>COX inhibitors,<br>number | PDA closure<br>after the 3rd course,<br>number |
| --- | --- | --- | --- | --- | --- | --- | --- | --- | --- | --- | --- | --- | --- | --- | --- |
| Szachnowicz<br>2026 | 2016–2023 | United<br>States | GA < 37 weeks | Ibuprofen or<br>Indomethacin | Intravenous | 97 | 50 | Median (IQR)<br>25.6 (24.3–27.0) | Median (IQR)<br>720 (600–900) | 97 | 35 | 25 | 7 |  |  |
| Gao<br>2026 | 2019–2024 | China | GA < 34 weeks | Ibuprofen | ND | 70 | 28 | Mean ± SD<br>29.4 ± 2.2 | Mean ± SD<br>1,179 ± 359 | 66 | 42 | 24 | 9 |  |  |
| Dani<br>2025 | 2018–2023 | Italy | GA < 29 weeks<br>(Subgroup:<br>GA < 25 weeks) | Ibuprofen | Intravenous | 107<br>(22–25 weeks, 32) | 52 | Mean ± SD<br>26.2 ± 1.5 | Mean ± SD<br>867 ± 226 | 92<br>(30) | 59<br>(13) | 27<br>(11) | 8<br>(3) |  |  |
| Cakir<br>2022 | 2017–2019 | Turkey | GA < 32 weeks and<br>Birth weight < 1,500 g | Ibuprofen | Intravenous | 169 | 79 | Median (IQR)<br>28 (2) | Median (IQR)<br>1,020 (290) | 169 | 123 | 46 | 23 | 23 | 15 |
| D'Amato<br>2021 | 2009–2018 | Italy | GA < 32 weeks and<br>Birth weight < 1,500 g | Ibuprofen | Intravenous | ELBW 54<br>VLBW 46 | ELBW 24<br>VLBW 21 | Mean ± SD<br>ELBW 27 ± 2<br>VLBW 29 ± 2 | Mean ± SD<br>ELBW 815 ± 135<br>VLBW 1,198 ± 149 | 62 | 52 | 10 | 6 |  |  |
| Meena<br>2020 | ND | India | GA < 37 weeks | Ibuprofen or<br>Indomethacin | Oral | Indomethacin 35<br>Ibuprofen 35 | Indomethacin 15<br>Ibuprofen 14 | Mean ± SD<br>Indomethacin 31.8 ± 2.3<br>Ibuprofen 31.4 ± 1.7 | Mean ± SD<br>Indomethacin 1,410 ± 320<br>Ibuprofen 1,340 ± 220 | 70 | 21 | 49 | 30 |  |  |
| Oboodi<br>2020 | 2016–2017 | Iran | GA < 37 weeks | Ibuprofen | Oral | 68 | 36 | Mean ± SD<br>31.1 ± 2.4 | Mean ± SD<br>1,353.6 ± 333.2 | 68 | 52 | 16 | 10 |  |  |
| Oh<br>2020 | 2010–2016 | Korea | GA < 30 weeks | Ibuprofen | Intravenous<br>or Oral | Responder 68<br>Non-responder 24 | Responder 38<br>Non-responder 14 | Median (range)<br>Responder 27.7 (23.6–29.9)<br>Non-responder 26.1 (23.3–29.3) | Median (range)<br>Responder 985 (410–1,560)<br>Non-responder 825 (460–1,160) | 92 | 68 | 19 | 5 |  |  |
| El-Farrash<br>2019 | 2015–2017 | Egypt | GA < 34 weeks | Ibuprofen | Oral | 30 | 14 | Mean ± SD<br>31.7 ± 2.0 | Mean ± SD<br>1,740 ± 470 | 30 | 12 | 18 | 12 |  |  |
| Kaur<br>2019 | 2014–2016 | Canada | GA < 32 weeks | Ibuprofen or<br>Indomethacin | Intravenous<br>or Oral | 183 | 84 | Median (IQR)<br>26 (25–28) | Mean ± SD<br>870 ± 313 | 183 | 120 | 62 | 30 | 12 | 6 |
| Dani<br>2019 | 2013–2017 | Italy | GA < 28 weeks | Ibuprofen | Intravenous | 23–24 weeks, 106<br>25–28 weeks, 405 | 23–24 weeks, 52<br>25–28 weeks, 207 | ND | Mean ± SD<br>23–24 weeks, 642 ± 116<br>25–28 weeks, 895 ± 214 | 363 | 221 | 135 | 47 |  |  |
| Olgun<br>2017 | 2008–2010 | Turkey | GA < 37 weeks | Ibuprofen | Oral | 97 | 52 | Mean ± SD<br>30.6 ± 3.4 | Mean ± SD<br>1,220 ± 480 | 94 | 67 | 27 | 11 | 14 | 5 |
| El-Mashad<br>2017 | 2012–2015 | Egypt | GA < 28 weeks and<br>Birth weight < 1,500 g | Ibuprofen or<br>Indomethacin | Intravenous | Indomethacin 100<br>Ibuprofen 100 | Indomethacin 60<br>Ibuprofen 80 | Mean ± SD<br>Indomethacin 26 ± 2.1<br>Ibuprofen 25 ± 2.1 | Mean ± SD<br>Indomethacin 1,100 ± 140<br>Ibuprofen 1,000 ± 120 | 200 | 158 | 41 | 12 |  |  |
| Sahin<br>2016 | 2011–2013 | Turkey | GA < 33 weeks | Ibuprofen | Oral | 60 | ND | Mean ± SD<br>29.2 ± 2.9 | ND | 60 | 49 | 11 | 9 |  |  |
| Pistulli<br>2014 | 2010–2012 | Albania | GA 28–32 weeks and<br>Birth weight < 2,000 g | Ibuprofen | Intravenous<br>or Oral | 68 | 37 | ND | ND | 68 | 53 | 15 | 12 |  |  |
| Chan<br>2014 | 2008–2011 | Hong Kong | GA < 37 weeks | Ibuprofen | Intravenous | 43 | 20 | Mean ± SD<br>28.8 ± 2.9 | Mean ± SD<br>1,148 ± 402 | 43 | 25 | 13 | 4 |  |  |
| Lago<br>2014 | 2008–2010 | Italy | GA < 32 weeks | Ibuprofen | Intravenous | 56 | 26 | Mean ± SD<br>27.4 ± 2.7 | Mean ± SD<br>1,027 ± 346 | 56 | 32 | 23 | 4 | 10 | 2 |
| Sivanandan<br>2013 | 2009–2011 | Canada | GA < 32 weeks<br>(Subgroup:<br>GA < 28 weeks) | Ibuprofen or<br>Indomethacin | Intravenous | Indomethacin 54<br>Ibuprofen 70 | Indomethacin 27<br>Ibuprofen 44 | Mean ± SD<br>Indomethacin 27 ± 2.3<br>Ibuprofen 27 ± 2.1 | Mean ± SD<br>Indomethacin 1,004 ± 272<br>Ibuprofen 992 ± 347 | 124<br>(81) | 71<br>(38) | 27<br>(26) | 9<br>(8) |  |  |
| Youn<br>2013 | 2009–2011 | Korea | Birth weight <1,500 g | Ibuprofen | Oral | 104 | ND | ND | ND | 104 | 41 | 58 | 34 |  |  |
| Adrouche-Amrani<br>2012 | 2004–2009 | United<br>States | Birth weight < 1,000 g | Ibuprofen or<br>Indomethacin | ND | 80 | 38 | Mean ± SD<br>25.2 ± 0.1 | Mean ± SD<br>753 ± 14 | 80 | 26 | 53 | 16 | 15 | 6 |
| van der Lugt<br>2012 | 2005–2011 | Netherlands | GA < 32 weeks | Ibuprofen | Intravenous | 164 | 81 | Median (range)<br>27.0 (21–31) | Median (range)<br>995 (525–2,041) | 164 | 109 | 43 | 24 | 11 | 6 |
| Moore<br>2012 | 1994–2005 | Canada | GA < 27 weeks | Indomethacin | ND | 196 | 106 | Median (range)<br>25 (23–26) | Median (range)<br>782 (355–1,300) | 167 | 60 | 88 | 28 | 21 | 3 |
| Erdeve<br>2012 | 2010–2011 | Turkey | GA < 28 weeks and<br>Birth weight < 1,000 g | Ibuprofen | Intravenous<br>or Oral | Intravenous ibuprofen 34<br>Oral ibuprofen 36 | Intravenous ibuprofen 13<br>Oral ibuprofen 20 | Mean ± SD<br>Intravenous ibuprofen 26.3 ± 1.3<br>Oral ibuprofen 26.4 ± 1.1 | Mean ± SD<br>Intravenous ibuprofen 872 ± 123<br>Oral ibuprofen 892 ± 117 | 70 | 44 | 26 | 24 |  |  |
| Lee<br>2012 | 2005–2010 | Taiwan | Birth weight < 1,500 g | Ibuprofen or<br>Indomethacin | Intravenous Indomethacin<br>or Oral Ibuprofen | Indomethacin 88<br>Ibuprofen 52 | Indomethacin 43<br>Ibuprofen 29 | Mean ± SD<br>Indomethacin 28.9 ± 2.2<br>Ibuprofen 28.8 ± 2.3 | Mean ± SD<br>Indomethacin 1,131 ± 223<br>Ibuprofen 1,130 ± 240 | 140 | 94 | 14 | 14 |  |  |
| Kushnir<br>2011 | 2005–2008 | United<br>States | GA < 37 weeks | Ibuprofen or<br>Indomethacin | Intravenous<br>or Oral | Indomethacin 165<br>Ibuprofen 185 | Indomethacin 82<br>Ibuprofen 108 | Indomethacin 27.7 (mean)<br>Ibuprofen 27.8 (mean) | Indomethacin 1,048 (mean)<br>Ibuprofen 1,083 (mean) | 344 | 197 | 125 | 33 | 56 | 10 |
| Tantawy<br>2011 | 2009 | Egypt | GA < 34 weeks | Ibuprofen | Oral | 100 | 48 | Mean ± SD<br>31.3 ± 1.3 | Mean ± SD<br>1,350 ± 271 | 100 | 76 | 19 | 9 |  |  |
| Gokmen<br>2011 | 2009–2010 | Turkey | GA < 32 weeks and<br>Birth weight < 1,500 g | Ibuprofen | Intravenous<br>or Oral | Intravenous 50<br>Oral 52 | Intravenous 29<br>Oral 22 | Mean ± SD<br>Intravenous 28.7 ± 2.1<br>Oral 28.5 ± 1.9 | Mean ± SD<br>Intravenous 1,205 ± 366<br>Oral 1,170 ± 297 | 102 | 74 | 28 | 26 |  |  |
| Richards<br>2009 | 1999–2006 | United<br>Kingdom | Birth weight <1,000g | Ibuprofen | ND | 160 | 75 | Mean ± SD<br>25.6 ± 1.4 | Mean ± SD<br>757 ± 127 | 160 | 72 | 80 | 32 |  |  |
| Vida<br>2009 | 2001–2007 | Italy | GA < 32 weeks | Ibuprofen | Intravenous | 201 | 99 | Median (IQR)<br>26 (25–28) | Median (IQR)<br>830 (650–1,015) | 201 | 92 | 100 | 51 | 24 | 12 |
| Sangem<br>2008 | 2000–2004 | United<br>States | GA < 34 weeks | Indomethacin | ND | 259 | ND | ND | ND | 259 | 198 | 61 | 26 | 23 | 10 |
| Yang<br>2008 | 2000–2003 | Singapore | Birth weight < 1,000 g | Indomethacin | Intravenous | 139 | 77 | Mean ± SD<br>26.3 ± 1.6 | Mean ± SD<br>807.3 ± 139.8 | 139 | 88 | 49 | 16 |  |  |
| Lee<br>2003 | 1998–2000 | Singapore | Birth weight < 1,500 g | Indomethacin | Intravenous | 69 | 37 | Mean ± SD<br>27.4 ± 2.6 | Mean ± SD<br>985 ± 269 | 69 | 47 | 17 | 6 |  |  |
| Lago<br>2002 | 1988–2000 | Italy | GA < 34 weeks<br>(Subgroup:<br>GA < 28 weeks) | Ibuprofen or<br>Indomethacin | Intravenous | Indomethacin 81<br>Ibuprofen 94 | Indomethacin 43<br>Ibuprofen 52 | Mean ± SD<br>Indomethacin 29 ± 3<br>Ibuprofen 28 ± 2 | Mean ± SD<br>Indomethacin 1,214 ± 427<br>Ibuprofen 1,126 ± 412 | 175<br>(83) | 123<br>(50) | 52<br>(32) | 25<br>(9) |  |  |

**Table S5.** Joanna Briggs Institute (JBI) Critical Appraisal Checklist for Case Series.

| Author,<br>Publication<br>Year | Clear Criteria<br>for inclusion | Reliable ways<br>for measuring<br>the condition | Valid methods<br>for identifying<br>inclusion | Consecutive<br>inclusion<br>of participants | Complete<br>inclusion<br>of participants | Clear reports on<br>the demographics<br>of the participants | Clear reports on<br>the clinical information<br>of the participants | Clear reports on<br>the outcomes or<br>follow-up results | Clear reports of<br>presenting sites or<br>demographic information | Appropriate<br>statistical<br>analysis | Overall<br>appraisal | Comments<br>(including reason<br>for exclusion) |
| --- | --- | --- | --- | --- | --- | --- | --- | --- | --- | --- | --- | --- |
| Szachnowicz<br>2026 | Unclear | No | Unclear | Yes | Yes | Yes | Yes | Unclear | Unclear | Yes | Include | N/A |
| Gao<br>2026 | Yes | Yes | Yes | Yes | Unclear | Yes | Yes | Unclear | Unclear | Unclear | Include | N/A |
| Dani<br>2025 | Yes | Yes | Yes | Yes | Yes | Yes | Yes | Yes | Yes | Yes | Include | N/A |
| Cakir<br>2022 | Unclear | Yes | Yes | Yes | Yes | Yes | Unclear | Unclear | Yes | Yes | Include | N/A |
| D'Amato<br>2021 | No | Yes | No | Yes | Yes | Yes | Yes | No | Yes | Yes | Include | N/A |
| Meena<br>2020 | No | Yes | Yes | Unclear | Unclear | Yes | Yes | Yes | No | Yes | Include | No description<br>on the study period |
| Oboodi<br>2020 | Yes | Yes | Yes | Yes | Yes | Yes | Yes | Yes | Yes | Yes | Include | N/A |
| Oh<br>2020 | Yes | Yes | Yes | Yes | Yes | Unclear | Yes | Yes | Yes | Yes | Include | N/A |
| El-Farrash<br>2019 | Yes | Yes | Yes | Yes | Yes | Yes | Yes | Unclear | Yes | Yes | Include | N/A |
| Kaur<br>2019 | Unclear | Unclear | Unclear | Yes | Yes | Yes | Yes | Yes | Yes | Yes | Include | N/A |
| Dani<br>2019 | Yes | Yes | Yes | Yes | Yes | Yes | Yes | Unclear | Yes | Yes | Include | N/A |
| Olgun<br>2017 | Yes | Yes | Yes | Yes | Yes | Yes | Yes | Yes | Yes | Yes | Include | N/A |
| El-Mashad<br>2017 | Yes | Yes | Yes | Yes | Yes | Yes | Yes | Unclear | Yes | Yes | Include | N/A |
| Sahin<br>2016 | Yes | Yes | Yes | Yes | Yes | No | No | Unclear | Yes | Yes | Include | N/A |
| Pistulli<br>2014 | Yes | Yes | Yes | Yes | Yes | No | No | No | Yes | Yes | Include | N/A |
| Chan<br>2014 | Yes | Unclear | Yes | Yes | Yes | Yes | Unclear | Unclear | Yes | Yes | Include | N/A |
| Lago<br>2014 | Yes | Yes | Yes | Yes | Yes | Yes | No | No | Yes | Yes | Include | N/A |
| Sivanandan<br>2013 | Yes | Yes | Yes | Yes | Yes | Yes | Yes | Yes | Yes | Yes | Include | N/A |
| Youn<br>2013 | Yes | Yes | Yes | Yes | Yes | No | No | Unclear | Yes | Yes | Include | N/A |
| Adrouche-Amrani<br>2012 | Yes | Yes | Yes | Yes | Yes | Yes | Yes | Yes | Yes | Yes | Include | N/A |
| van der Lugt<br>2012 | Yes | Yes | Yes | Yes | Yes | Yes | Yes | Yes | Yes | Yes | Include | N/A |
| Moore<br>2012 | Yes | Yes | Yes | Yes | Yes | Yes | Yes | Yes | Yes | Yes | Include | N/A |
| Erdeve<br>2012 | Yes | Yes | Yes | Yes | Yes | Yes | Yes | Unclear | Yes | Yes | Include | N/A |
| Lee<br>2012 | Yes | Yes | Yes | Yes | Yes | Yes | Yes | Unclear | Yes | Yes | Include | N/A |
| Kushnir<br>2011 | Yes | Yes | Unclear | Yes | Yes | Yes | Yes | Unclear | Yes | Yes | Include | N/A |
| Tantawy<br>2011 | Yes | Yes | Yes | Yes | Yes | Unclear | Unclear | Unclear | Yes | Yes | Include | N/A |
| Gokmen<br>2011 | Yes | Yes | Yes | Yes | Yes | Yes | Yes | Unclear | Yes | Yes | Include | N/A |
| Richards<br>2009 | Yes | Yes | Yes | Yes | Yes | Yes | Yes | Yes | Yes | Yes | Include | N/A |
| Vida<br>2009 | Yes | Yes | Yes | Yes | Yes | Yes | Yes | Yes | Yes | Yes | Include | N/A |
| Sangem<br>2008 | Yes | Yes | Yes | Yes | Yes | Yes | Yes | Yes | Yes | Yes | Include | N/A |
| Yang<br>2008 | Yes | Unclear | Yes | Yes | Yes | Yes | Yes | No | Yes | Yes | Include | N/A |
| Lee<br>2003 | Yes | Yes | Yes | Yes | Yes | Yes | Yes | Yes | Yes | Yes | Include | N/A |
| Lago<br>2002 | Unclear | Unclear | Unclear | Yes | Yes | Yes | Yes | Unclear | Yes | Yes | Include | N/A |

**Table S6.** Multilevel meta-regression analysis of trends across treatment courses.

| Category | Population/Subgroup | *n* | Odds Ratio [95% CI] | *p*-value (Trend) |
| --- | --- | --- | --- | --- |
| Overall population | All preterm infants | 33 | 0.56 [0.50–0.62] | < 0.001 |
| GA and birth weight | EPT/ELBW infants | 10 | 0.50 [0.41–0.61] | < 0.001 |
|  | Preterm infants from non-overlapping cohorts | 26 | 0.58 [0.51–0.66] | < 0.001 |
| Drug type | Ibuprofen | 27 | 0.59 [0.51–0.67] | < 0.001 |
|  | Indomethacin | 10 | 0.44 [0.36–0.54] | < 0.001 |
| Route | Intravenous route | 16 | 0.51 [0.44–0.60] | < 0.001 |
|  | Oral route | 8 | 0.99 [0.73–1.35] | 0.947 |

*n*, number of studies; CI, confidence interval; GA, gestational age; EPT, extremely preterm; ELBW, extremely low birth weight. The odds ratio represents the relative increase in the likelihood of PDA closure per additional treatment course, estimated using multilevel mixed-effects meta-regression accounting for within-study correlations.

**Table S7.**
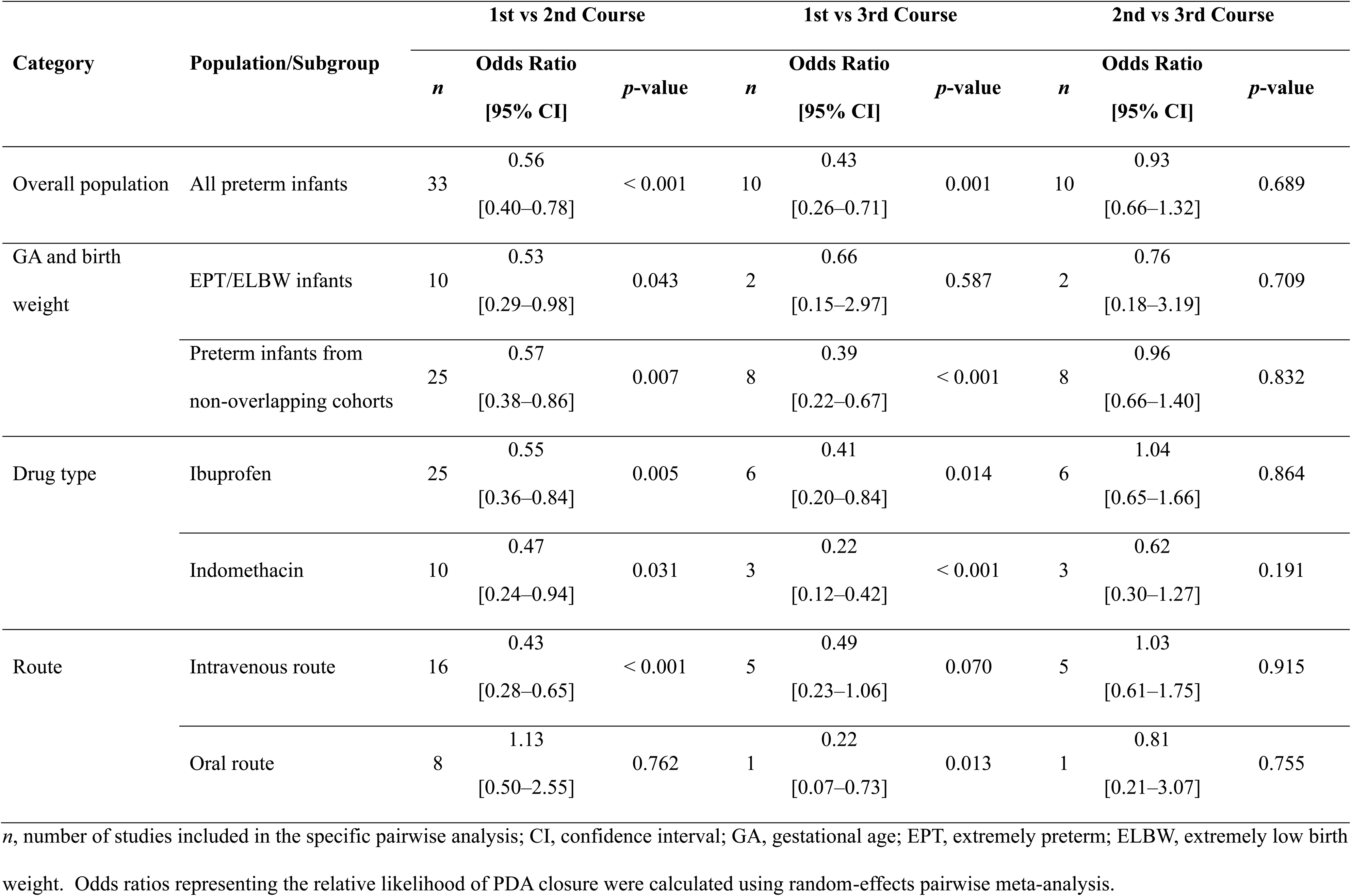
Pairwise comparisons of PDA closure rates across treatment courses.

**Table S8.**
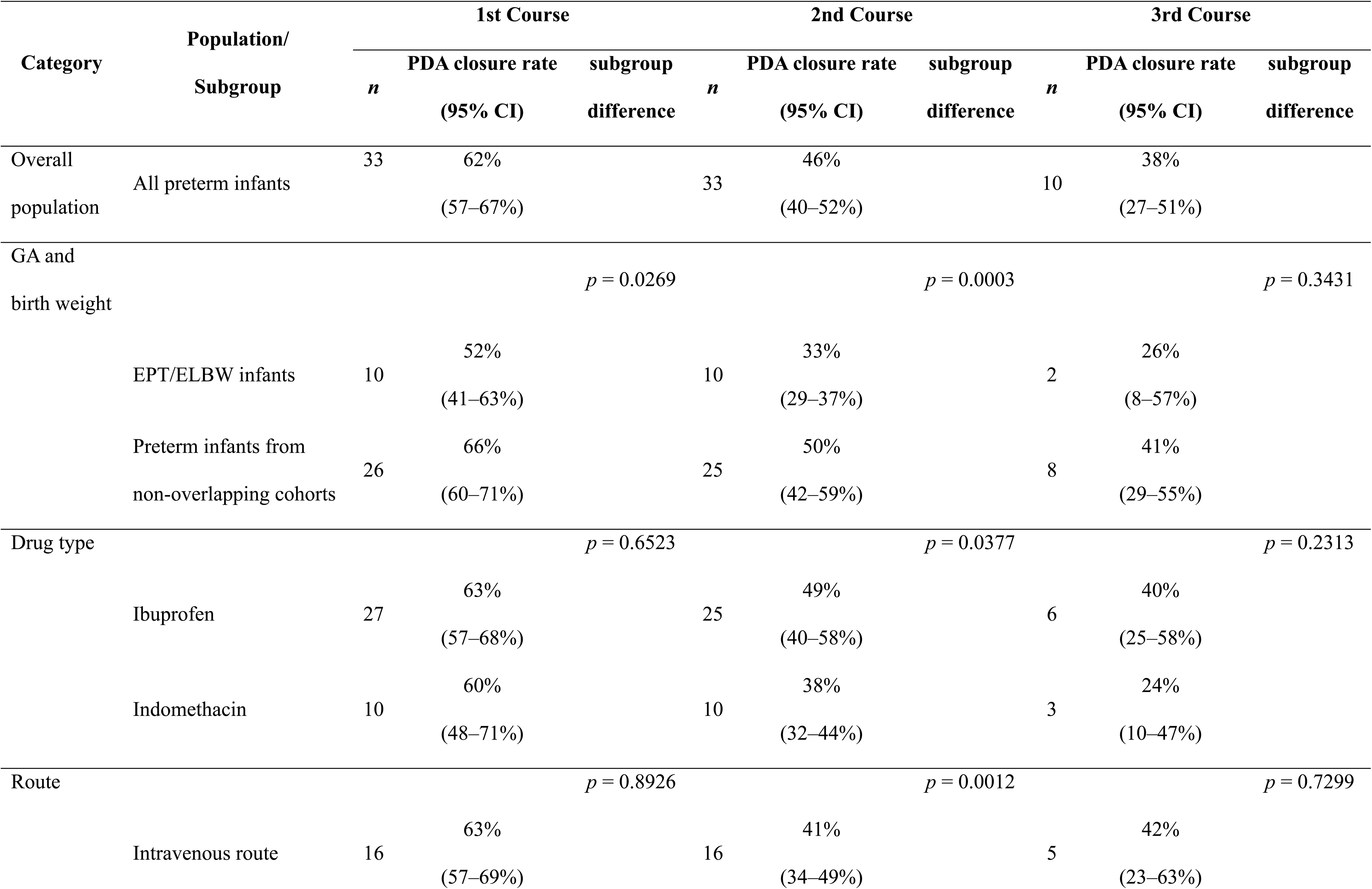
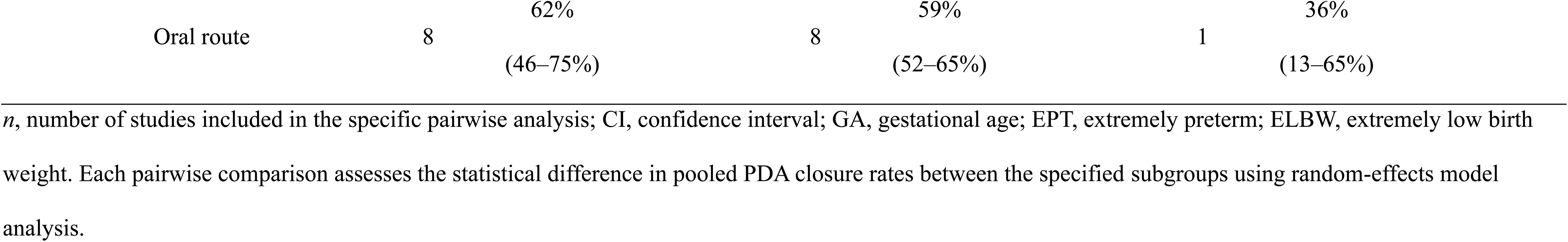
Subgroup comparisons of PDA closure rates by treatment courses.

| Category | Population/<br>Subgroup | 1st Course |  |  | 2nd Course |  |  | 3rd Course |  |  |
| --- | --- | --- | --- | --- | --- | --- | --- | --- | --- | --- |
|  |  | *n* | PDA closure rate | subgroup | *n* | PDA closure rate | subgroup | *n* | PDA closure rate | subgroup |
|  |  |  | (95% CI) | difference |  | (95% CI) | difference |  | (95% CI) | difference |
| Overall | All preterm infants | 33 | 62% |  | 33 | 46% |  | 10 | 38% |  |
| population |  |  | (57–67%) |  |  | (40–52%) |  |  | (27–51%) |  |
| GA and |  |  |  | *p* = 0.0269 |  |  | *p* = 0.0003 |  |  | *p* = 0.3431 |
| birth weight | EPT/ELBW infants | 10 | 52% |  | 10 | 33% |  | 2 | 26% |  |
|  |  |  | (41–63%) |  |  | (29–37%) |  |  | (8–57%) |  |
|  | Preterm infants from | 26 | 66% |  | 25 | 50% |  | 8 | 41% |  |
|  | non-overlapping cohorts |  | (60–71%) |  |  | (42–59%) |  |  | (29–55%) |  |
| Drug type |  |  |  | *p* = 0.6523 |  |  | *p* = 0.0377 |  |  | *p* = 0.2313 |
|  | Ibuprofen | 27 | 63% |  | 25 | 49% |  | 6 | 40% |  |
|  |  |  | (57–68%) |  |  | (40–58%) |  |  | (25–58%) |  |
|  | Indomethacin | 10 | 60% |  | 10 | 38% |  | 3 | 24% |  |
|  |  |  | (48–71%) |  |  | (32–44%) |  |  | (10–47%) |  |
| Route |  |  |  | *p* = 0.8926 |  |  | *p* = 0.0012 |  |  | *p* = 0.7299 |
|  | Intravenous route | 16 | 63% |  | 16 | 41% |  | 5 | 42% |  |
|  |  |  | (57–69%) |  |  | (34–49%) |  |  | (23–63%) |  |

|  |  |  |  |  |  |  |
| --- | --- | --- | --- | --- | --- | --- |
| Oral route | 8 | 62%<br>(46–75%) | 8 | 59%<br>(52–65%) | 1 | 36%<br>(13–65%) |

*n*, number of studies included in the specific pairwise analysis; CI, confidence interval; GA, gestational age; EPT, extremely preterm; ELBW, extremely low birth weight. Each pairwise comparison assesses the statistical difference in pooled PDA closure rates between the specified subgroups using random-effects model analysis.

**Table S9.**
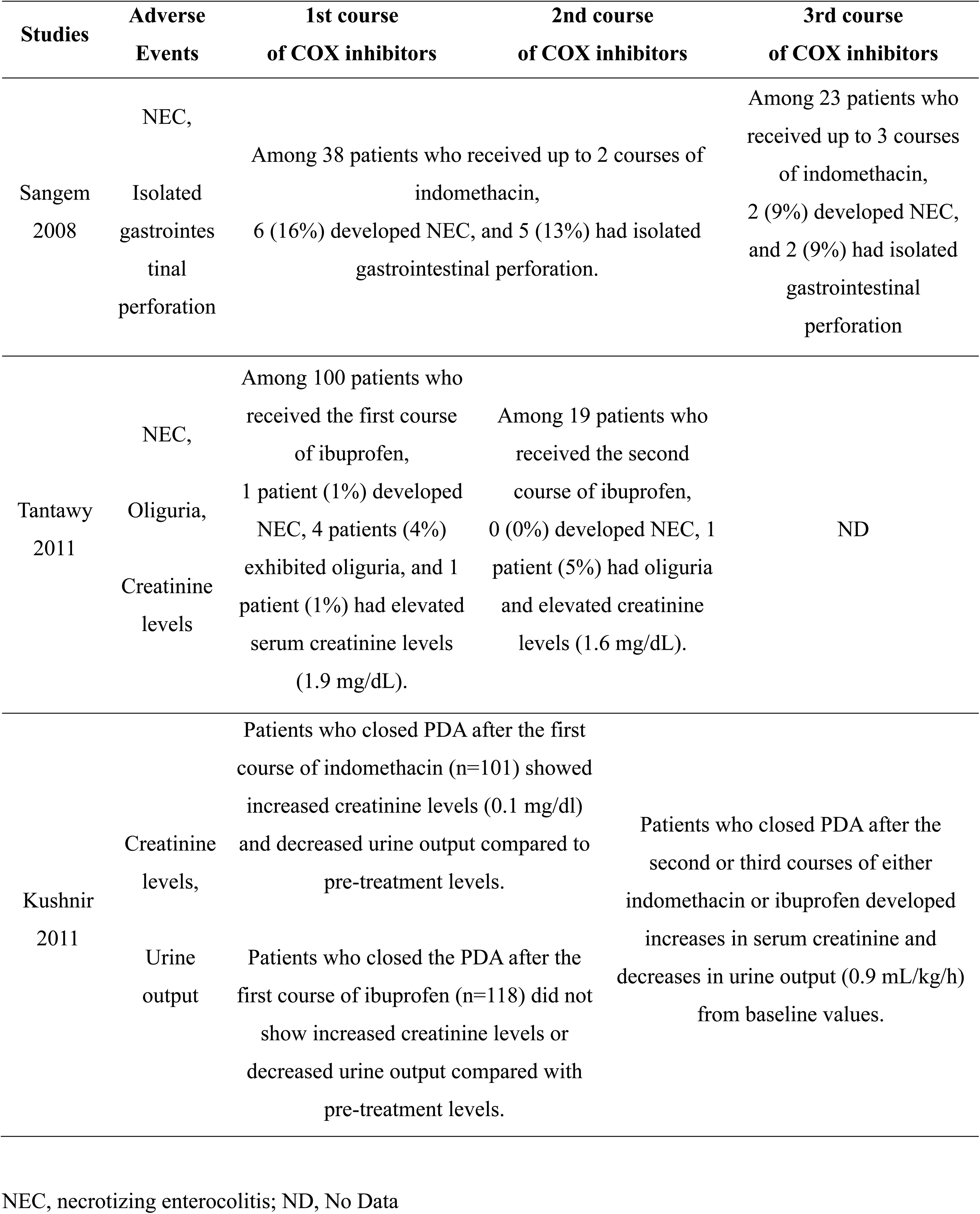
Summary of adverse events after each course of COX inhibitor treatment.

**Table S10.** GRADE Evidence Profile.

| **Outcome** | **Related Figures or Tables** | **Number of Studies** | **Study Design** | **Risk of Bias** | **Inconsistency** | **Indirectness** | **Imprecision** | **Publication Bias** | **Certainty** |
| --- | --- | --- | --- | --- | --- | --- | --- | --- | --- |
| PDA closure,<br>All preterm infants | Figs. 2, 3, S9,<br>Tables S6, S7, S8 | 33 | Case series extracted from<br>RCTs and non-RCTs | Not serious | Not serious | Not serious | Not serious | Not likely | Low |
| PDA closure,<br>EPT/ELBW infants | Figs. 4, 5, S9,<br>Tables S6, S7, S8 | 10 | Case series extracted from<br>RCTs and non-RCTs | Not serious | Not serious | Not serious | Not serious | Not likely | Low |
| PDA closure,<br>Ibuprofen | Figs. S1, S2, S10,<br>Tables S6, S7, S8 | 27 | Case series extracted from<br>RCTs and non-RCTs | Not serious | Not serious | Not serious | Not serious | Not likely | Low |
| PDA closure,<br>Indomethacin | Figs. S3, S4, S10,<br>Tables S6, S7, S8 | 10 | Case series extracted from<br>RCTs and non-RCTs | Not serious | Not serious | Not serious | Not serious | Not likely | Low |
| PDA closure,<br>Intravenous | Figs. S5, S6, S11,<br>Tables S6, S7, S8 | 15 | Case series extracted from<br>RCTs and non-RCTs | Not serious | Not serious | Not serious | Not serious | Not likely | Low |
| PDA closure,<br>Oral | Figs. S7, S8, S11,<br>Tables S6, S7, S8 | 8 | Case series extracted from<br>RCTs and non-RCTs | Not serious | Not serious | Not serious | Not serious | Not likely | Low |
| Renal dysfunction<br>(Indomethacin vs Ibuprofen) | Table S9 | 1 (Kushnir 2011) | Observational Study | Not serious | N/A | Not serious | Serious | N/A | Very low |
| Renal dysfunction<br>(1st course vs 2nd course) | Table S9 | 1 (Tantawy 2011) | Case series | Not serious | N/A | Not serious | Serious | N/A | Very low |
| Gastrointestinal complications<br>(1st course vs 2nd course) | Table S9 | 1 (Tantawy 2011) | Case series | Not serious | N/A | Not serious | Serious | N/A | Very low |
| Gastrointestinal complications<br>(2nd course vs 3rd course) | Table S9 | 1 (Sangem 2008) | Case series | Not serious | N/A | Not serious | Serious | N/A | Very low |

